# Impaired HA-specific T follicular helper cell and antibody responses to influenza vaccination are linked to inflammation in humans

**DOI:** 10.1101/2021.04.07.21255038

**Authors:** Danika L Hill, Silvia Innocentin, Jiong Wang, Eddie A James, James C Lee, William W Kwok, Martin Zand, Edward J Carr, Michelle A Linterman

## Abstract

Antibody production following vaccination can provide protective immunity to subsequent infection from pathogens such as influenza. However, circumstances where antibody formation is impaired after vaccination, such as in older people, require us to better understand the cellular and molecular mechanisms that underpin successful vaccination in order to improve vaccine design for at risk groups. Here, by studying the breadth of anti-hemagglutinin (HA) IgG, serum cytokines, and B and T cell responses by flow cytometry before and after influenza vaccination, we show that formation of circulating T follicular helper cells (cTfh) cells are the best predictor of high titre antibody responses. Using MHC class II tetramers we demonstrate that HA-specific cTfh cells can derived from pre-existing memory CD4^+^ T cells and have a diverse TCR repertoire. In older people, the differentiation of HA-specific cells into cTfh cells was impaired. This age-dependent defect in cTfh cell formation was not due to a contraction of the TCR repertoire, but rather was linked with an increased inflammatory gene signature in cTfh cells. Together this suggests that strategies that temporarily dampen inflammation at the time of vaccination may be a viable strategy to boost optimal antibody generation upon immunisation of older people.

**One sentence summary:** Antibody production upon vaccination requires antigen-specific cTfh cells whose formation is suppressed by pro-inflammatory cytokine signalling.

## Introduction

Vaccination is an excellent intervention to limit the morbidity and mortality caused by infectious disease. Yet, despite their success, most vaccines are not completely effective, and efficacy varies significantly between different vaccines. The seasonal influenza vaccine needs to be administered each year in order to provide protection against the most prevalent circulating influenza strains, but its efficacy typically ranges from 40-80% even when the vaccine is antigenically matched to circulating viruses. This inefficacy contributes to the millions of severe influenza cases and hundreds of thousands of deaths globally (*1*), which could be potentially prevented by a more effective vaccine.

The reasons that the seasonal influenza vaccine provides protection in some individuals, but not others, have yet to be fully established. Antibodies against the influenza surface glycoprotein haemagglutinin (HA) are capable of limiting infection, and anti-HA antibody titres and inhibitory activity are the most commonly used correlate of protection (*2*). The human antibody response to influenza vaccination is highly variable, but what causes this inter-individual variation is not well understood. Twin studies estimate that genetics can account for less than 20% of the variation in antibody responses to influenza vaccination, implicating non-heritable factors as key contributing influences (*3*). Age, sex, chronic viral infections, and non-communicable diseases have all been reported to influence antibody titre following vaccination (*4-9*), but how these various factors impact immune responses to vaccination have yet to be fully unravelled.

The generation of protective humoral immunity is supported by CD4^+^ helper T cells (*10*), which, like neutralising antibodies, are correlates of protection for influenza infection (*11*). The majority of work on human T cell responses to influenza vaccination has focussed on T helper type 1 cells, largely because of the relative ease of detecting antigen-specific cytokine-secreting cells upon ex vivo peptide restimulation. However, this approach fails to identify T cell types, such as Tfh cells, that do not readily secrete cytokines(*12, 13*), and therefore our understanding of how the human CD4 T cell response is linked with high titre antibody responses upon vaccination is limited. Here, we use MHC class-II tetramers (*14, 15*) to directly assess helper T cell responses to the seasonal influenza vaccine. We find that differentiation of antigen-specific circulating T follicular helper (cTfh) cells, but not the total number of HA-specific CD4^+^ T cells, is correlated with high titre antibody production upon vaccination. HA-specific cTfh cells are clonally expanded from memory cells present pre-vaccination, and share a transcriptional profile with human lymph node Tfh cells. Further, we find that in older people there is a specific defect in the formation of cTfh cells upon vaccination. Interestingly, this was not explained by limited T cell receptor diversity of the responding T cells, as is commonly proposed as the cause of poor T cell responses in older people (*16, 17*). Rather, poor cTfh and antibody responses correlated with an enhanced inflammatory gene signature. Together, this implicates cTfh cells as key mediators of antigen-specific immunity and suggests that vaccine strategies that limit, rather than enhance, the inflammation associated with ageing may be more successful in older individuals.

## Results

### Hemagglutinin-specific CD4^+^ T cells expand and differentiate in response to seasonal influenza vaccination

In order to track influenza HA-specific CD4 T cells directly *ex vivo* we recruited two cohorts of healthy UK individuals between 18-36 years old across two norther hemisphere influenza seasons in 2014/2015 (n=16, median 30.5 years old) and 2016/2017 (n=21, median 25 years old) carrying either *HLADR*0701* or *HLADR*1101* alleles. Blood samples were collected at baseline (d0), seven days (d7), and 42 days after seasonal trivalent influenza vaccination (TIV). A total of 53 distinct variables were measured at two or more time-points (n=147 total measures, Fig 1A, Table S1, Figure S1-3). IgG levels increased against all measured HA proteins from the vaccine influenza strains at d7 and d42 (Fig 1B). We analysed post-vaccination time-points (d7 and d42) compared to baseline to identify which immunological parameters were altered by vaccination (Fig. 1C). From our panel of 32 HA proteins, IgG titres to 31 (96%) were altered at day 42 relative to baseline (d0), with the greatest fold changes were observed for HA strains contained in the TIV. These data indicate that vaccine-induced IgG responses were able to partially cross-react across multiple influenza strains (Fig. 1C). The increase in anti-HA antibody titre was coupled with an increase in hemagglutination inhibitory antibodies to A.Cali09, the one influenza A strain contained in the TIV that was shared across the two cohorts (Fig. 1C). Our analysis of 8 cytokines by Luminex identified that 4 cytokines were upregulated (CXCL13, BCMA, APRIL and Osteopontin) and 2 were downregulated (BAFF and TWEAK) at d7 post-vaccination (Fig. 1C, S4). We did not detect alterations in the frequency of any B cell subsets either cohort by flow cytometry, however the frequency of cTfh cells (CD45RA^-^CXCR5^+^PD1^++^) was increased on d7, a population that we and others have previously shown share transcriptional and clonal similarity with germinal centre Tfh cells (*18-21*). Furthermore, the expression of ICOS on cTfh cells was increased on d7, confirming the cTfh population is an activated effector population that forms in response to vaccination (Fig. 1C, S4).

**Figure 1:**
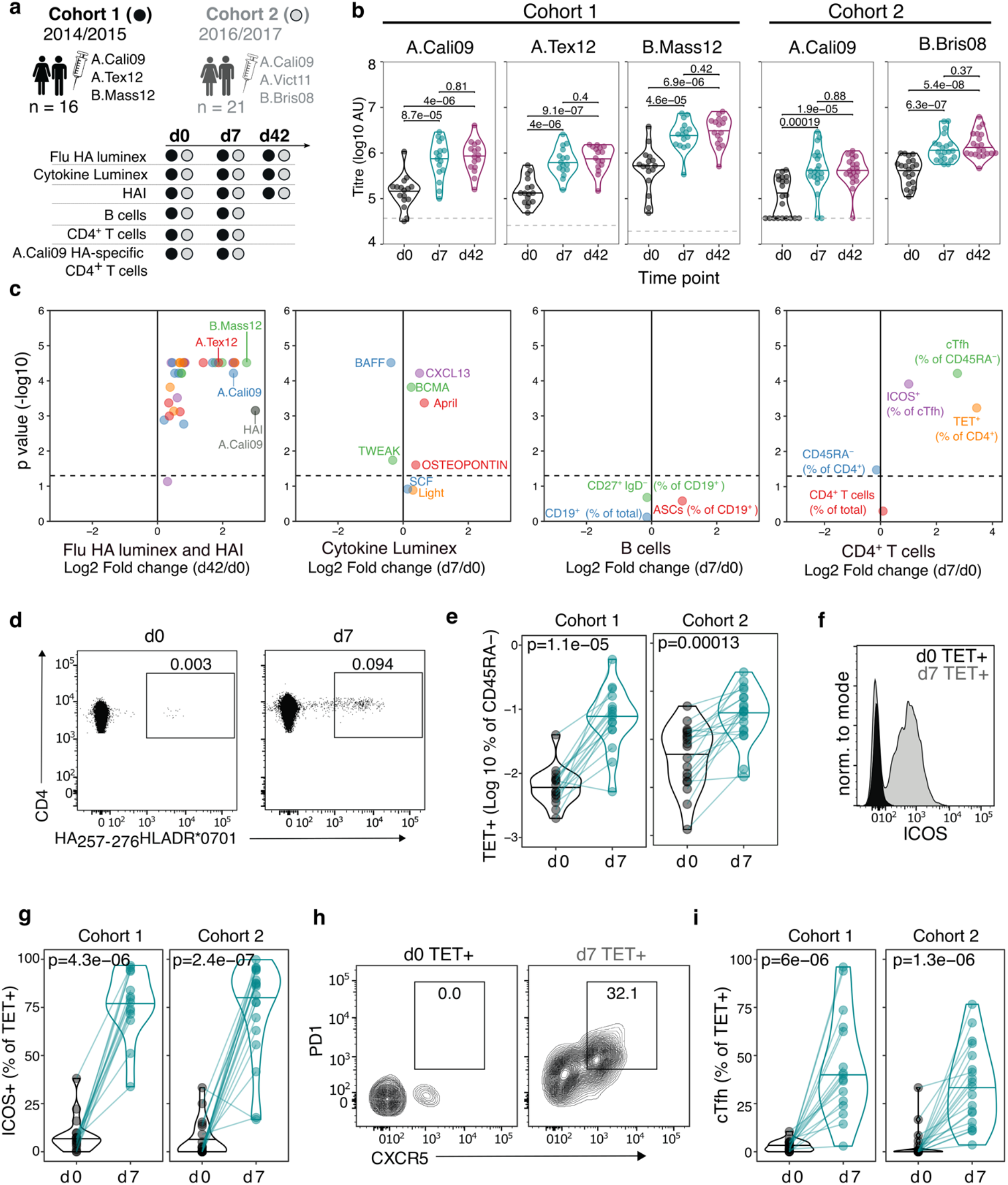
Robust hemagglutinin-specific CD4^+^ T cells response to seasonal influenza vaccination. a) Overview of cohort characteristics, vaccination strains and immune variables measured before (d0), seven days (d7) and 42 days (d42) after seasonal influenza vaccination. b) IgG responses to Haemagglutinin proteins from vaccine influenza strains measured by Luminex. Dashed line indicates limit of detection in Luminex assay. c) Log_2_ fold change versus -log10 FDR-adjusted p-value of Flu HA Luminex, Cytokine Luminex, B cell and CD4^+^ T cell immunophenotyping parameters before and after vaccination at indicated time-points for Cohort 1. Dashed line represents the p-value cut off at FDR-adjusted p=0.05. d) Representative flow cytometry plots of HA_257-276_HLADR*0701 staining on CD4^+^CD45RA^-^ cells, gate corresponds to HA-specific TET^+^ T cells, and e) the frequency of HA-specific TET^+^ T cells among all CD4^+^CD45RA^-^ cells on d0 compared to d7. f) Representative ICOS staining, and g) the percentage of ICOS^+^ cells among HA-specific TET^+^ T cells on d0 and d7. h) CXCR5 and PD1 staining on HA-specific TET^+^ T cells on d0 and d7, gate corresponds to ‘TET^+^cTfh’ T cells, and i) the percentage of cTfh cells among HA-specific TET^+^ T cells on d0 and d7 for each cohort. In parts d-i; Cohort 1 n= 16; Cohort 2 n = 19. Paired p-values determined using Wilcoxon signed rank-test.

To study HA-specific CD4^+^ T cell responses, we focussed our analysis on the A.Cali09 strain as this was included in both seasons’ vaccine formulations, and used MHC Class II tetramers of HLADR*0701 and HLADR*1101 loaded with A.Cali09 HA peptides to identify antigen-specific T cells (*15*). Tetramer binding antigen-experienced CD4^+^CD45RA^-^ (Tet^+^) T cells showed the largest fold-change increase after vaccination of any parameter measured (Fig. 1C). Tet^+^ cells were detected in all individuals before vaccination and expanded a median of 5-11-fold between d0 and d7 in both cohorts (Fig. 1D, E). These antigen-specific T cells had upregulated ICOS after immunisation, indicating that they have been activated by vaccination (Fig. 1F, G). In addition, one third of HA-specific T cells upregulated the Tfh markers CXCR5 and PD1 seven days after immunisation (Fig. 1H, I), the majority of which expressed CXCR3 (Fig. S5), consistent with the “Th1” skew in the total CD4^+^ T cell (*15*) and cTfh cell response to influenza vaccination (*22*). Therefore, seasonal influenza vaccination increases anti-HA IgG titres, induces a cTfh response, and promotes the expansion and differentiation of HA-specific CD4^+^ T cells.

### Circulating HA-specific Tfh cells correlate with vaccine IgG response

The majority of successful vaccines provide protection against re-infection through the production of antibodies. The development of antibody secreting plasma cells requires a concerted effort of multiple cell types of the immune system, which have been investigated in detail in mice, but not well in humans. Therefore we sought to determine which immune parameters were linked with A.Cali09 IgG titre six weeks post-vaccination (Fig. 2A-D). Pre-existing antibody titres have been linked with diminished responses to subsequent vaccination (*23-25*), but while we observed a slight negative correlation at d0 in support of this, the relationship was not statistically significant for any of the Flu HA Luminex or A.Cali09 HAI titres (Fig. 2B). In contrast, the IgG responses to a range of HA proteins from different influenza strains correlated strongly with A.Cali09 IgG (d42-d0), indicating that those individuals with large vaccine-induced IgG responses also developed cross-reactive antibodies against multiple strains (Fig. 2B). Changes between d0 and d7 in serum BCMA was positively correlated, and BAFF negatively correlated with A.Cali09 IgG (d42-d0) responses, in both cohorts (Fig. 2C, E). The frequency of B cell subsets, including antibody secreting cells at d7 was not associated with day 42 A.Cali09 IgG (d42-d0), whereas cTfh cell frequency correlated with antibodies in both cohorts (Fig. 2D, E). A reproducible positive correlation was observed between A.Cali09 IgG (d42-d0) and HA-specific cTfh cells (Fig. 2D, E), but not for total Tet^+^ CD4^+^ T cells (Fig. 2D). This indicates that the differentiation of antigen-specific Tfh cells is more relevant for antibody responses than the overall frequency of antigen-specific helper T cells.

**Figure 2:**
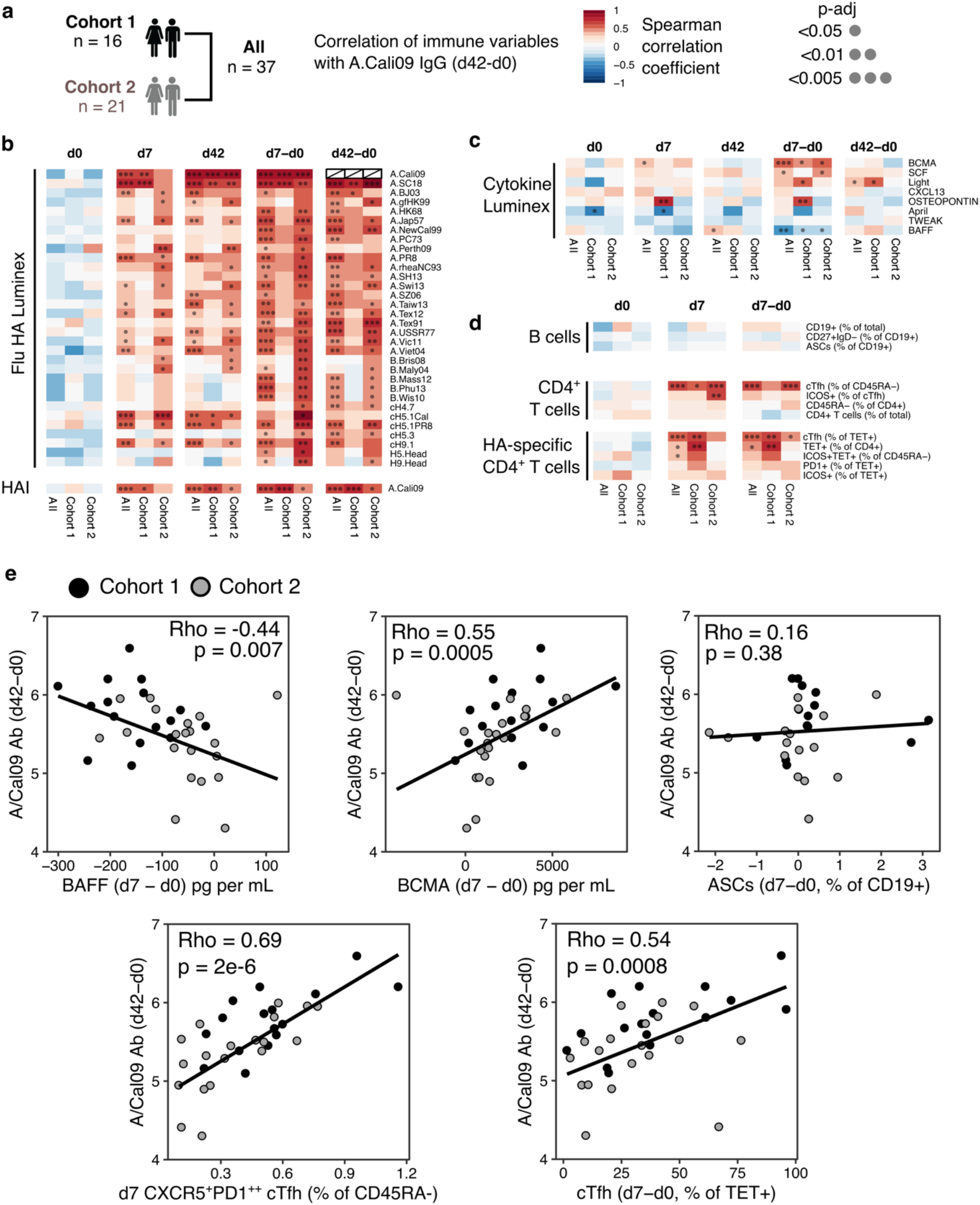
Circulating HA-specific Tfh cells correlate with vaccine IgG response. a) Overview of correlation analysis between A.Cali09 IgG response at day 42 (minus d0 baseline titer, d42-d0) and immune variables for Cohort 1, Cohort 2 and both cohorts combined (All). b) Correlations for Flu HA Luminex IgG and hemagglutination inhibition (HAI) titres at d0, d7 and d42, and at d7 and d42 after subtracting each individuals d0 baseline value (d7-d0, d42-d0). c) Correlations for serum cytokines measured by Luminex at d0, d7 and d42, and at d7 and d42 after subtracting each individuals d0 baseline value (d7-d0, d42-d0). d) Correlations for B cell, CD4^+^ T cell and HA-specific CD4^+^ T cell variables at d0, d7 and d7 after subtracting each individuals d0 baseline value (d7-d0). e) Correlation between vaccine-induced A.Cali09 IgG at d42 with selected immune parameters in both Cohort 1 and Cohort 2 (n=37). Dot color corresponds to the cohort (black = Cohort 1, grey = Cohort 2). Coefficient (Rho) and p-value determined using Spearman’s correlation, and line represents linear regression fit.

### HA-specific cTfh response to influenza vaccination includes recalled and public TCR clonotypes

Our results demonstrated that HA-specific cTfh cells are correlated with the antibody response to seasonal influenza vaccination, and so we next investigated how vaccination influences the T cell receptor (TCR) repertoire and transcriptional signatures of HA-specific cTfh cells and their precursors. We sort-purified and RNA sequenced tetramer-binding CD4^+^ T cells (Tet^+^ cells) from d0 and Tet^+^ cTfh cells from d7 (Fig. 3A, S3, S6), and retrieved a total of 1405 and 2085 TCRβ clonotypes at d0 and d7, respectively. Expanded clones were observed on d7 (Fig. 3B), resulting in a decrease in the diversity of the TCR repertoire at d7 relative to d0 (Fig. 3C), and an increase in the number of co-dominant TCRβ clonotypes within the HA-specific TCR repertoire of each individual (Fig. 3D). Analysis of paired samples that were sequenced at both d0 and d7 demonstrated that TCRβ clonotypes were recalled from the d0 memory CD4^+^ T cell pool into the cTfh cell response at d7 in 10 out of 15 individuals (Fig. 3E, F). Recalled TCRβ clones represented a median of 25% of the overall TCR repertoire at d7 (Fig. 3G), and were more likely to have been more abundant at d0 compared to clones only detected at d0 (Fig. 3H). Next, we sought to compare pre- and post-vaccination TCR repertoires between individuals. Identical TCRβ sequences between 2 or more individuals, or ‘public clonotypes’, were present in 18 out 20 cTfh Tet^+^ samples at d7, and represented a median of 10% of the total response (total of 32 distinct public TCR sequences, Fig. 3I-J). In contrast, only 4 public clonotypes were shared across 7 individuals at d0. There was a tendency for public clonotypes to preferentially use particular *TCRBV* and *TCRBJ* gene combinations (Fig. 3K). These results demonstrate that the vaccine-induced cTfh response to A.Cali09 involves reactivation of existing memory CD4^+^ T cells, and that common responses between individuals are a prevalent part of the cTfh cell response.

**Figure 3:**
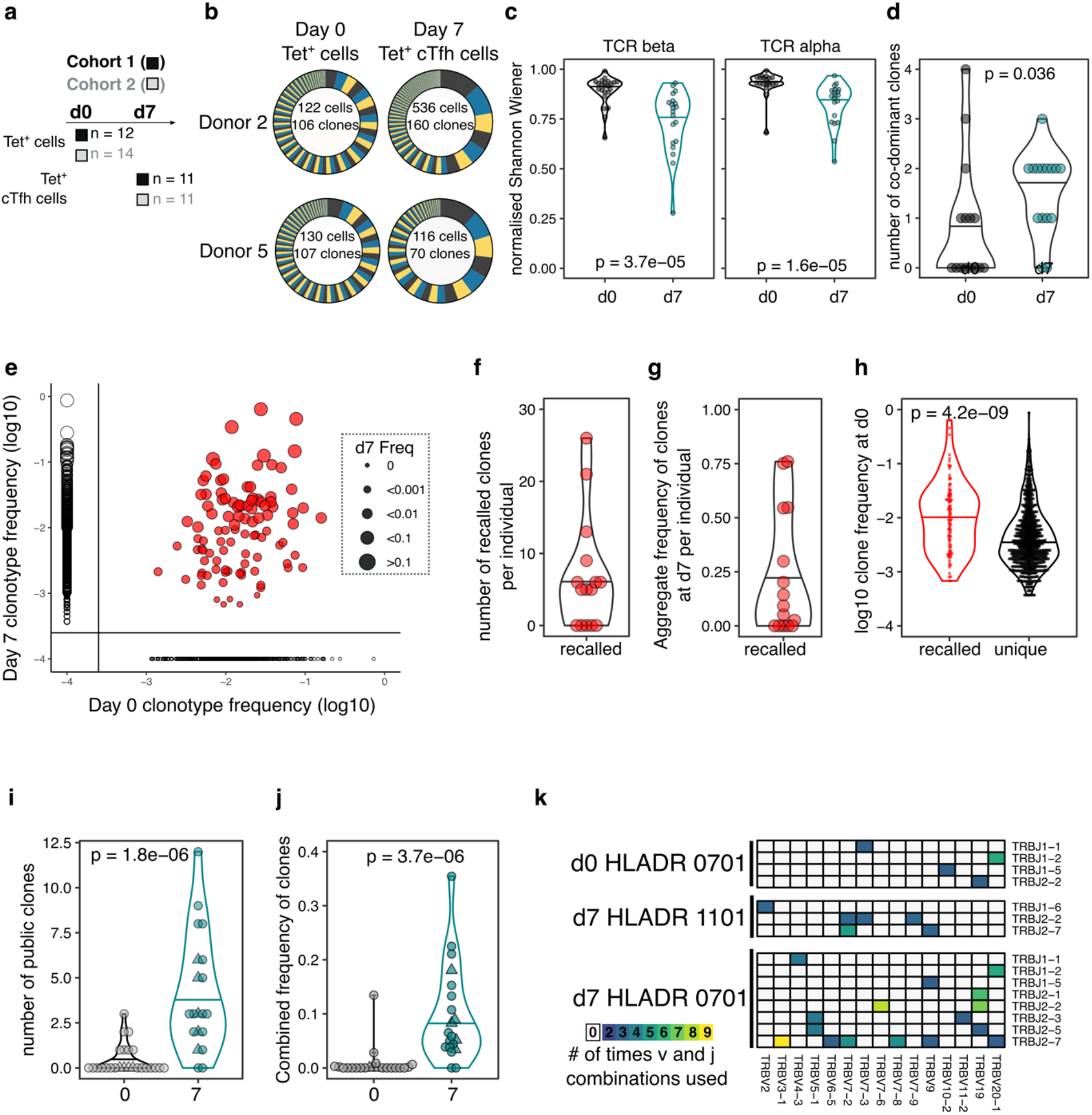
HA-specific cTfh response to flu vaccination includes recalled and public TCR clonotypes. a) Overview of cell types and sample sizes that were sequenced d0 and d7 at each cohort. d0 median = 45 cells (6-258); d7 median = 88 cells (5-1000). b) Representative pie charts of the proportions unique T cell receptor (TCR) β-chain clonotypes for participants 408S and 425L at d0 and d7. Inset numbers indicate the number of cells sequenced and number of unique TCRβ clonal sequences retrieved. c) Normalised Shannon Wiener index of TCRβ and TCRα repertoire diversity at d0 TET^+^ T cells and d7 TET^+^ cTfh cells, for both cohorts combined. d) The number of dominant TCRβ clones (frequency of >10%) for each individual in d0 and d7 sequenced cells for both cohorts combined. e) TCRβ clonotype frequencies at d0 and d7 in combined dataset of 15 individuals with paired d0 and d7 samples (1772 clones in total). Each dot represents a clonotype, size corresponds to d7 frequency. Red color indicates “recalled” clones that were measured at both time-points, and solid lines indicate where clones are only detected at a single time-point. f) The number of ‘recalled’ TCRβ clones per individual (present at both d0 and d7), and g) the aggregate frequency of d7 TET^+^ cTfh TCRβ clones that were recalled for each individual (n= 15). h) The log10 d0 frequency of each “recalled” or “unique” (present at d0 only) clone (n = 102 recalled, n = 663 unique clones). i) The number and j) combined frequency of public clonotypes per individual at each time point, cohorts 1 and 2 combined (d0 n= 25, d7 n = 20). Public clonotypes were defined as TCRβ sequences with identical v, d, j genes and CDR3 amino acid sequence shared between at two or more samples from the same time point. k) TRBJ and TRBV gene usage among the 26 HA-specific public clonotypes, separated by HLADR genotype and time-point. All p-values were calculated using a Mann-Whitney *U* test.

### Vaccination-induced cTfh cells share a common transcriptional signature with lymph node Tfh cells

We sought to determine the transcriptional profile of HA-specific cTfh (Tet^+^) cells, and to determine to what extent these cTfh Tet^+^ cells acquired the transcriptional signature of *bona fide* human lymph node germinal centre Tfh cells. HA-specific cTfh cells that form seven days after vaccination clustered distinctly from and their d0 Tet^+^ precursors cells by principal component analysis (PC1) for both cohorts (Fig. 4A). 684 genes were differentially expressed in Tet^+^ cTfh cells compared to d0 Tet^+^ cells across both cohorts (DESeq2 Log_2_FC > 0.5, FDR p-value <0.1, Data File S1). This gene signature was compared to a 1179 gene signature generated by comparing germinal centre Tfh cells (CXCR5^+^PD1^+++^) to non-Tfh antigen-experienced CD4^+^ T cells from human inguinal lymph node (*20*) (Data File S2). 147 of the differentially expressed genes in Tet^+^ cTfh cells shared the same expression pattern with genes up or downregulated in germinal centre Tfh signature, termed “Tfh genes”, and included upregulation of key Tfh transcription factors *MAF* and *TOX2*, and downregulation of several cytokine and chemokine receptors such as *IL7R, IL2RA*, and *CCR6* (Fig. 4B, C, Data File S3). The remaining 537 genes, termed ‘Vaccination genes’, were modulated after influenza vaccination but not differentially expressed in our human lymph node Tfh cell samples. This list included the downregulation of *CXCR4*, a chemokine receptor that is important for lymph node homing consistent with the blood localisation of the cTfh cells (*26*), and upregulation of the interferon-gamma inducible gene *GBP2*. This gene-level analysis indicated that Tet^+^ cTfh cells share features of a transcriptional profile with lymph node Tfh cells, while also expressing genes specific either to the Th1-skewed response to influenza vaccination or to circulation in the blood.

**Figure 4:**
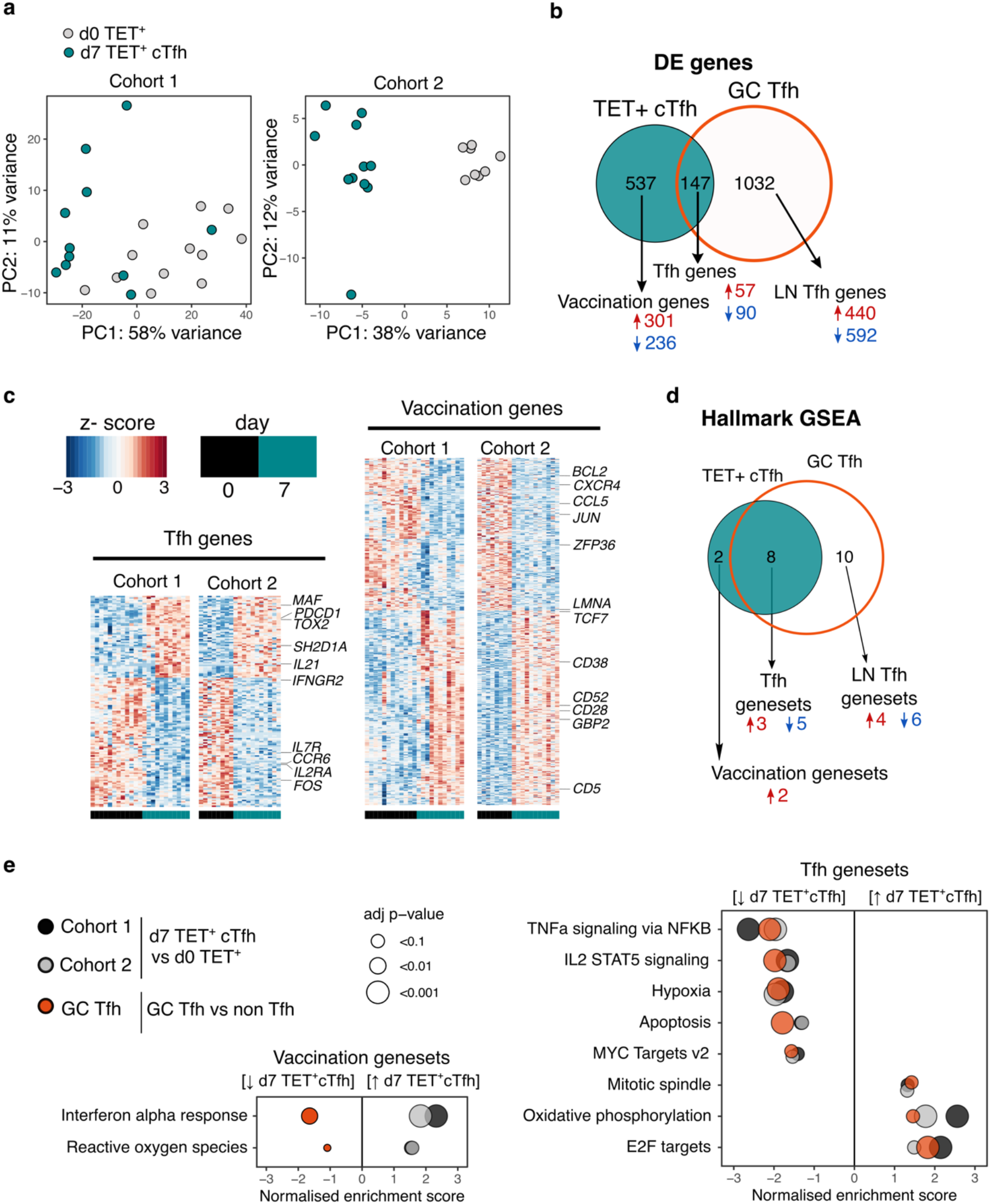
Vaccination induced circulating Tfh cells share a common transcriptional signature with lymph node Tfh cells. a) Principal component analysis of 1000 genes with the largest variance in sequenced cells from d0 TET^+^ cells (shown in grey) and d7 TET^+^ cTfh cells (shown in green), for each cohort separately. b) Venn diagram of the 684 significantly differentially expressed (DE) genes in d7 TET^+^cTfh cells (relative to d0) in both cohorts, and the overlap with a signature of human lymph node germinal centre (GC) Tfh cells where genes had the same direction of expression. DE genes determined using DESeq2, and had adjusted p-value <0.1 and fold change of 2. Up and down regulated genes represented by red and blue colours, respectively. c) Heatmaps of gene signatures determined in (b) for each cohort, with representative genes labelled. d) Venn diagram of the 10 consistently enriched Hallmarks pathways in d7 TET^+^cTfh cells relative to d0 TET+ cells in both cohorts, and the overlap with positively or negatively enriched gene sets in LN GC Tfh cells compared to non-Tfh CD4^+^ cells. Significant pathways were determined using gene set enrichment analysis (GSEA) and had adj p-value <0.1 and normalised enrichment score of >1 or <-1. Up and down regulated pathways are represented by arrows and red and blue colours, respectively. e) Bubble plots showing the normalised enrichment scores for significantly enriched pathways for d7 TET^+^ cTfh cells versus d0 TET^+^ cells in each cohort, and LN GC Tfh cells vs non-Tfh cells, with positive and negative scores indicating positive and negative enrichment, respectively, in TET+ Tfh and/or GC Tfh compared to their non-Tfh comparator. Circle colour represents the type of comparison, and size represents the adjusted p-value.

To gain a clearer understanding of what molecular pathways were modulated during the differentiation into Tfh cells after vaccination at a transcriptional level, we performed gene set enrichment analysis using the Hallmarks gene sets (Fig. 4D)(*27*). Ten gene sets were differentially modulated in d7 Tet^+^ Tfh cells compared to d0 in both cohorts, 8 (80%) of which were also enriched in lymph node germinal centre Tfh cells (Fig. 4D). These ‘Tfh genesets’ included the downregulation several cytokine pathways including IL-2 signalling, a cytokine known to inhibit Tfh differentiation (*28, 29*), and the upregulation of oxidative phosphorylation, a metabolic process known to be elevated in Tfh cells (Fig. 4E) (*30, 31*). Enrichment for the interferon alpha response and reactive oxygen species pathways were not shared with lymph node Tfh cells, and may reflect the acute response to inactivated virus in the vaccine to which the lymph node Tfh cells were not exposed. Overall, our analysis demonstrates that seasonal influenza vaccination recalls antigen-specific memory CD4^+^ T cells and promotes their differentiation into Tfh cells with similarity to lymph node GC Tfh cells.

### Impaired anti-HA IgG responses in older individuals correlates with failure to fully acquire the immune trajectory of younger individuals

We defined the correlates of influenza vaccine antibody responses in 18-36 year olds to be serum concentrations of BAFF and BCMA, and the frequency of both polyclonal and HA-specific cTfh cells. Next we wanted to investigate how the immune response to seasonal influenza vaccination was impacted in older individuals, a group where the influenza vaccine is less efficacious (*32, 33*). We compared our cohorts of 18-36 year olds with that of individuals over 65 years old in the same vaccination years (Fig. 5A, cohort 1 median 69 years old, cohort 2 median 73.43 years old), and observed that older individuals showed lower HA-specific IgG responses compared to 18-36 year olds for all the vaccine strains measured (Fig. 5B). To investigate whether ageing broadly influenced immune status pre- and post-vaccination, we applied a diffusion pseudotime algorithm to 23 antibody, cytokine or immunophenotyping variables measured at d0 and d7 for both age-groups and cohorts (Table S1). The immune profiles formed a single continuous spectrum, with the pseudotime ‘vaccination trajectory’ beginning with d0 samples and ending with d7 samples from 18-36 year old samples (Fig. 5C). There was no pre-vaccination difference in trajectory values between the age-groups, indicating that ageing did not impact the baseline status for the immune parameters involved in the vaccination response (Fig. 5D). In contrast, over 65 year olds failed to progress along the vaccination trajectory to the same extent as their 18-36 year olds counterparts (Fig. 5D), indicating a failure to appropriately respond to vaccination. 11 immune parameters correlated with the vaccination trajectory values, including antibody responses to the vaccination strains, cTfh cells, HA-specific T cells and diminished serum BAFF (Fig. 5E). These results indicate that in over 65 year olds there is a failure to fully engage the adaptive immune system in response to vaccination compared to younger individuals.

**Figure 5:**
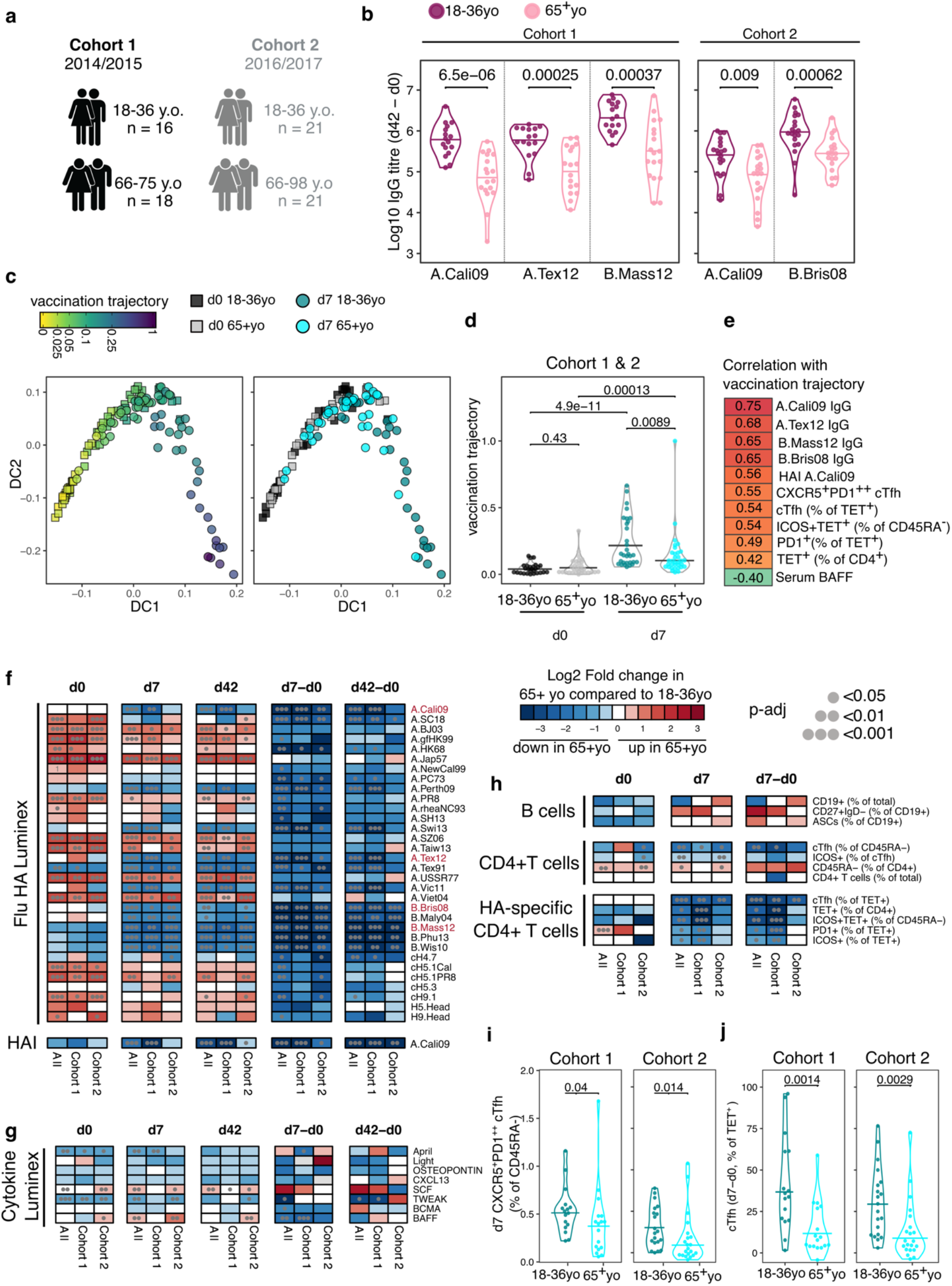
Impaired vaccination immune trajectory in older individuals. a) Overview of age-groups and sample sizes for each cohort. b) IgG responses to HA proteins from vaccine influenza strains measured by Luminex for each age-group. c) Diffusion map dimensionality reduction of 122 samples from both cohorts combined using scaled values for 23 immune parameters and the diffusion-pseudotime algorithm (d0 18-36yo n= 27; d0 65+yo n= 32; d7 18-36yo n= 30; d7 65+yo n=33). Each dot represents a sample, shape represents time point (d0 = squares, d7 = circles), and colour either the pseudotime ‘vaccination trajectory’ output value or age-group category. Diffusion components (DC) 1 and 2 shown. d) Vaccination trajectory values for sample in each age category from both cohorts combined, with p-values calculated using Dunn’s post hoc test. e) Spearman correlation coefficients for the 11 parameters that significantly correlated with the vaccination trajectory variable (padj<0.05). f-h) Heatmap of FDR-adjusted p-values from Mann-Whitney U test comparing immune parameters between age-groups for Cohort 1, Cohort 2 and both cohorts combined (All), at d0, d7 and d42, and at d7 and d42 after subtracting each individuals d0 baseline value (d7-d0, d42-d0) for f) Flu HA Luminex and HAI, g) Cytokine Luminex, h) B cells, CD4^+^ T cells and HA-specific CD4^+^ T cells. Colour corresponds to p-value and the direction of change. i) The percentage of CXCR5^+^PD1^++^ cTfh cells and j) TET^+^ cTfh cells for each age-group and each cohort, with p-values calculated by Mann-Whitney *U* test (Cohort 1 18-36yo n= 17, 65+yo n=17; Cohort 2 18-36yo n=20, 65+yo n=21).

In order to identify which immune parameters may explain the age-related difference in vaccination trajectory, we compared the 53 immune parameters between age-groups before and after vaccination. Older individuals had higher titres of IgG to several HA proteins from different influenza A strains at d0, however no difference was seen in IgG or HAI titre levels for the influenza strains in the vaccines administered to our cohorts (Fig. 5F) suggesting that age-dependent differences in pre-existing antibodies to the vaccine strain does not account for the diminished response. After immunisation, over 65 year olds showed lower vaccine induced IgG responses for numerous HA strains, consistent with the lower IgG responses to the vaccination strains and diminished generation of cross-reactive IgG to other influenza strains (Fig. 5F). For serum cytokines, we observed consistently lower baseline April and TWEAK concentrations, and higher d42 SCF levels in over 65 year olds across both cohorts (Fig. 5G, S7), but there was no age-dependent difference in BAFF or BCMA. No age-related differences in circulating B cell populations were observed pre- or post-vaccination, and no consistent differences were seen for CD4^+^ T cell subsets pre-vaccination, including for the frequency of d0 HA-specific CD4^+^ cells (Fig. 5H, S7). At d7, the frequency of polyclonal cTfh cells and HA-specific Tet^+^ cTfh cells were the only CD4^+^ T cell subsets consistently reduced in older individuals across both cohorts, with the strongest effect on antigen-specific cTfh cells (Fig. 5H-J). There was no consistent difference in the total d7 Tet^+^ HA-specific T cell population with age for both cohorts (Fig. 5H, S7), which suggests that the poor vaccine antibody responses in older individuals is impacted by impaired cTfh cell differentiation (Fig. 5J) rather than size of the vaccine-specific T cell pool.

### TCR repertoire diversity in HA-specific cTfh cells is comparable in younger and older individuals

Poor T cell responses to vaccination have been previously attributed to a contraction of the naïve CD4^+^ T cell population and restriction of the TCR repertoire with age (*16, 17*), although most human studies have examined the whole repertoire, rather than focusing on vaccine-specific T cells. Our ability to track vaccine specific CD4^+^ T cells enables direct assessment of the repertoire of responding cells, and therefore we sought to determine whether there were ageing-related differences in the clonal diversity of pre or post vaccination of HA-specific CD4^+^ T cells (Fig. 6A). The pre-vaccination diversity is important as our data from young donors demonstrates that a large contribution to the d7 response originates from memory CD4^+^ T cells present prior to vaccination. In pre-vaccination HA-specific Tet^+^ cells there was a slight reduction in TCR diversity in over 65 year olds compared to younger people, although this was only statistically significant in one cohort (Fig. 6B). However, after vaccination no difference was seen in the TCR diversity of the resulting d7 Tet^+^cTfh population between age groups (Fig. 6B), despite the diminished frequency of this population in older individuals. Likewise, no age-related difference in TCR diversity was observed for the total pool of vaccine-specific Tet^+^ population at day 7 (Fig. 6C). As we had observed in younger donors, TCRβ clones were present in both the d0 Tet^+^ and d7 Tet^+^ cTfh populations. There was a trend towards a lower absolute number of recalled clones in older individuals (Fig. 6D), in line with the lower numbers of d7 sequenced cells due to the reduced cTfh population frequency (Fig. S6). However, the proportion of the d7 population that consisted of recalled clones was not impacted by age (Fig. 6E), indicative of equivalent contribution from pre-existing memory cells into the resulting cTfh population between younger and older donors. The affinity of TCR for antigen shapes Tfh differentiation and Tfh vs. Th1 cell fate after influenza vaccination (*34-36*), and therefore we sought to determine if the age-related decline in Tet^+^ cTfh differentiation could be explained by an age-dependent skew in the TCR repertoire away from Tfh differentiation. We examined the TCRβ clones that were present in both the d7 Tet^+^ and d7 Tet^+^ cTfh populations, and observed strong correlations between clone frequency between the two populations irrespective of age-groups, suggesting that there was no restriction in the TCR repertoire able to give rise to Tfh cells to the HA peptides studied here (Fig. 6F). Together, our data indicate that TCR diversity is not a limiting factor for the Tfh cell response to seasonal influenza vaccination in older individuals, and suggests that the age-associated reduction in cTfh cell frequency is due to other cell-intrinsic or environmental effects on Tfh cell differentiation or survival.

**Figure 6:**
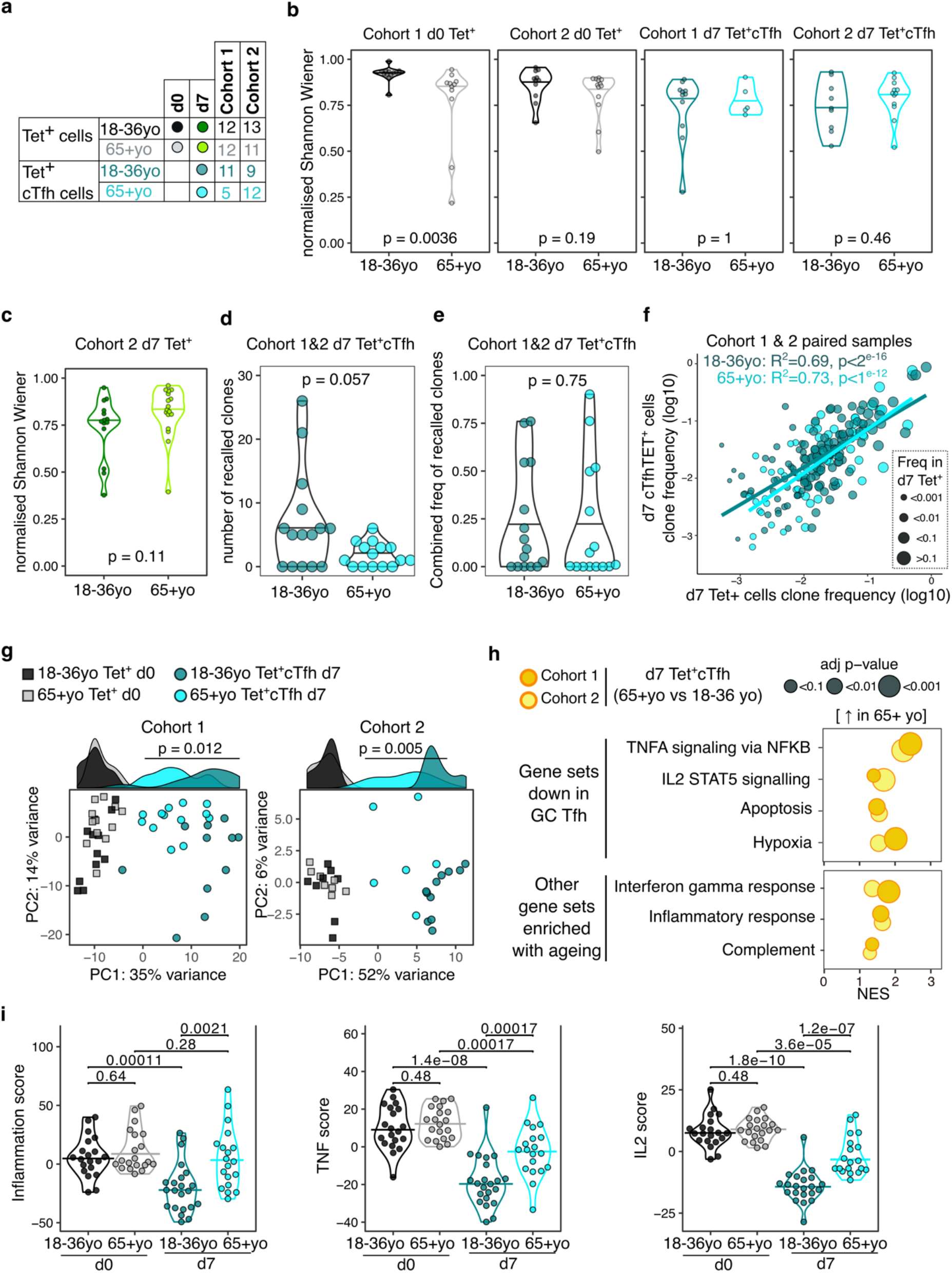
Impaired induction of Tfh transcriptional programs and aberrant inflammatory signatures in HA-specific cTfh cells from older individuals. a) Overview of sample numbers and cell types sequenced at d0 and d7 for each cohort and each age-group. Dark and light green dots indicate d7 Tet^+^ cells sequenced from Cohort 2 only. b) Normalised Shannon Wiener diversity index of TCRβ clonotypes for each cohort, time-point and cell type. Each dot represents a sequenced sample. c) Normalised Shannon Wiener diversity index of TCRβ clonotypes in d7 Tet^+^ cells from Cohort 2. d) The number of TCRβ clones per individual in d7 Tet^+^ cTfh cells recalled from d0 Tet^+^ cells, and e) combined frequency of the recalled clones among d7 Tet^+^cTfh cells for each age-group (n= 15 in each age-group, both cohorts combined. P-values calculated using Mann-Whitney U test. f) Frequency of TCRβ clones present in paired samples of d7 Tet^+^ and d7 Tet^+^cTfh cells. Each dot represents a clonotype, dot color indicates age-group, and dot size corresponds the frequency in d7 Tet^+^ cells. Lines represent linear regression, and statistics determined using Pearson correlation. g) Principal component (PC) analysis of the 684 genes differentially expressed (DE) between d0 and d7 in samples from 18-36 year olds applied to d0 and d7 samples from both age-groups and cohorts. Histograms show the distribution of PC1 values for each group, with p-value calculated between age-groups for d7 samples by Mann Whitney U test. h) Bubble plots of 7 Hallmarks pathways that are upregulated in d7 Tet^+^cTfh cells in samples from 65+year-old compared to 18-36-year-old individuals, sub-divided by which pathways were previously identified as negatively enriched in lymph node germinal centre Tfh cells. Positive scores indicate positive enrichment in Tet^+^cTfh cells from older donors.Circle colour represents the cohort, size represents the adjusted p-value. i) Inflammation, TNF and IL2 gene signature scores in HA-specific T cells at different time-points and age-groups, cohorts 1 and 2 combined (d0 18-36yo n= 20, d0 65+yo n = 20; d7 18-36yo n= 22; 65+yo n = 19). p-value calculated using Dunn’s post-hoc test.

### HA-specific cTfh cells from older individuals fail to induce Tfh transcriptional signatures and display aberrant inflammatory signatures

We investigated the transcriptome of HA-specific cTfh cells from older individuals to determine if we could identify pathways that could explain the poor Tfh cell differentiation in ageing. With supervised principal component analysis using the 684 genes that are differentially expressed in 18-36 year olds d7 cTfh cells compared to d0, we observed that d7 cTfh cells from over 65 year olds were clustered between d0 and d7 samples from younger donors for PC1 in both cohorts (Fig. 6G). Indeed, of the 425 genes that were consistently DE between HA-specific cells from d0 and d7 in older donors from both cohorts, only 170 (40%) were part of the younger d7 signature (Data file S5). Importantly, we did not observe any consistent age-related gene expression differences in pre-vaccination d0 cells, indicating that ageing is not associated with transcriptional changes in resting HA-specific memory cells. These data indicate that cTfh cells from older individuals failed to acquire the full gene signature seen in Tfh cells from younger people.

To further resolve these age-related transcriptional differences, we performed enrichment analysis with the Hallmarks genesets and observed 7 gene sets that were consistently positively enriched in d7 HA-specific Tet^+^cTfh cells from over 65 year old compared to 18-36 year old individuals (Fig. 6H). Four of these elevated genesets (TNFa signalling via NFKB; IL2 STAT5 signalling; Apoptosis; Hypoxia) we previously identified as negatively enriched in Tet^+^cTfh from 18-36 year old individuals compared to d0 Tet^+^ cells. This indicates that these genes are normally downregulated as HA-specific CD4^+^ T cells differentiate into Tfh cells in response to vaccination, and that this process appears dysregulated in older individuals. The remaining three enriched genesets (Interferon gamma response; Inflammatory response; Complement) are suggestive of heightened pro-inflammatory responses in cTfh cells from older individuals, and included enrichment for *IL6, CCL2* and *CCL5*. No pathways were altered by ageing in pre-vaccination d0 Tet^+^ cells, indicating that the enrichment for numerous inflammatory pathways in Tfh cells from older people occurred as part of the response to vaccination, rather than being a generalisable feature of CD4^+^ biology in ageing. To further investigate the upregulation of inflammation and cytokine signalling in cTfh cells from older donors, we used the leading-edge output from the gene set enrichment results to curate 3 non-overlapping gene lists of ‘TNF’, ‘IL2’ and ‘Inflammation’ (Table S2). We then calculated 3 gene list scores by generating gene z-scores for each cohort, and then summing the z-scores for the genes in each of the gene lists for each sample, before and after vaccination. For all three gene signatures, while there was no pre-vaccination difference between the age-groups, d7 cTfh cells from older donors failed to downregulate the expression of these genes to the same extent as cTfh cells from 18-36 year olds (Fig. 6I). In summary, our transcriptional analysis indicated that in older individuals, cTfh cells display evidence of inflammatory cytokine signalling that is not typical of Tfh cells in younger people.

### Vaccine induced transcriptional signatures of inflammation, TNF and IL-2 signalling negatively correlate with Tfh differentiation and antibody responses

We sought to determine whether the down-regulation of the inflammatory, IL-2 and TNF pathways in Tet^+^cTfh cells was linked to Tfh cell differentiation and antibody responses after vaccination. Irrespective of age, the three gene scores were negatively correlated with the Tet^+^ cTfh cell frequency (Fig. 7A), and the vaccine-induced A.Cali09 IgG response (Fig. 7B). Our findings suggest that the presence of pro-inflammatory cytokine signalling in cTfh cells induced by vaccination negatively impacts optimal vaccine responses. In order to determine whether the observed negative correlation between the three inflammatory gene signatures and IgG response to seasonal influenza vaccination could be replicated in other cohorts, we analysed PBMC microarray data and A.Cali09 anti-HA antibodies from an independent cohort of 50 adults aged 18-86 year olds from the U.S.A (*8*). Consistent with our cohorts, we did not detect any age-related difference in gene signatures present prior to vaccination, but the gene signatures induced by vaccination d7 (relative to d0) in PBMCs were negatively correlated with IgG production at d28 (Fig. 7C). This demonstrated that expression of genes associated with inflammation, TNF, and IL-2 signalling in blood is negatively associated with impaired IgG responses to seasonal influenza vaccination, irrespective of age. These data suggest that generation of a high-titre antibody response to vaccination requires the cytokine milieu in secondary lymphoid tissues to be carefully controlled to limit persistent pro-inflammatory cytokine signalling during Tfh differentiation.

**Figure 7:**
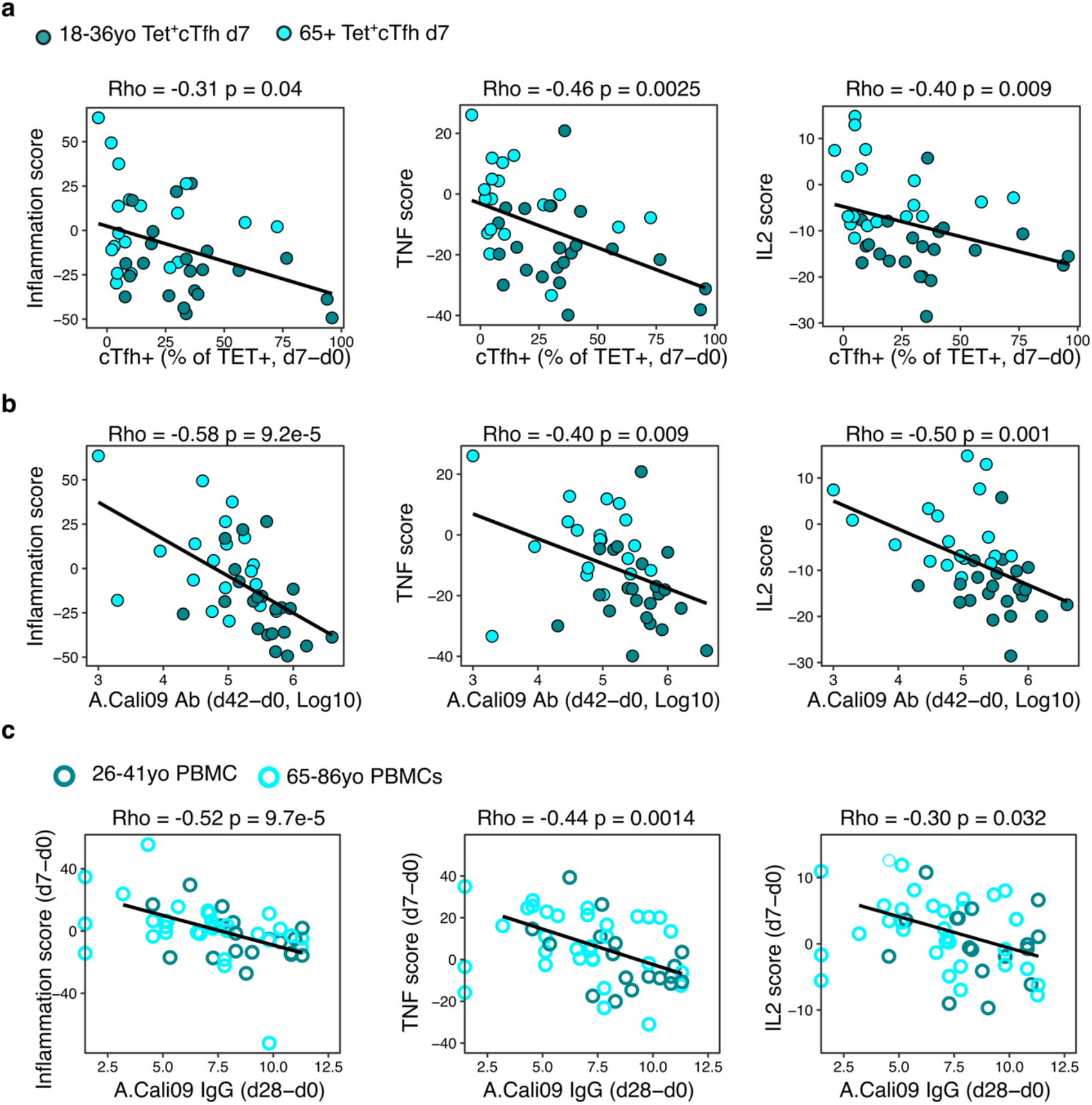
Gene signatures of TNF, IL-2, and inflammation associate with impaired antibody and Tfh cell responses to influenza vaccination. a) Correlation between cTfh Tet^+^ cells and Inflammation, TNF or IL2 gene signatures scores in d7 Tet^+^cTfh cells. b) Correlation between A.Cali09 IgG titre (d42-d0) and Inflammation, TNF or IL2 gene signatures scores in d7 Tet^+^cTfh cells. c) Correlation between A.Cali09 IgG titre (d28-d0) and Inflammation, TNF or IL2 gene signatures scores determined from microarray data of peripheral blood mononuclear cells on d0 or d7 after seasonal influenza vaccination from publicly available datasets (n= 50 total; 26-41yo n=18, 66-86yo = 32)(*8*). Correlation coefficients and p-values calculated using Spearman’s Correlation coefficient. Solid line represents linear regression fit. Color corresponds to age-group (Green = younger people; aqua = older people).

## Discussion

The formation of virus neutralising antibodies after influenza vaccination provides protections against subsequent infection, yet the cellular and molecular pathways that support a high-titre antibody response in humans remain incompletely defined. Here a systems immunology approach was used to determine which immune parameters are associated with antibody formation upon vaccination. We used MHC class II tetramers to track the formation and differentiation of haemagglutinin-specific CD4^+^ T cells after vaccination, ensuring the specificity of the CD4^+^ T cell response can be accurately matched to the A.Cali09 antibody response. HA-specific cTfh cell frequency was strongly correlated with anti-A.Cali09 antibodies six weeks after vaccination, and there was not a reproducible relationship between total HA-specific CD4^+^ T cells and antibody titre, highlighting the importance of antigen-specific Tfh cell differentiation to support humoral immunity. TCR repertoire analysis showed that HA-specific cTfh cells formed from pre-existing memory cells in both younger and older adults, but their differentiation was reduced in older people. Interestingly, there was no difference in TCR repertoire diversity in cTfh cells in ageing, indicating that a contraction of the TCR repertoire with ageing is unlikely to be the cause of poor Tfh cell differentiation in older people. The defective cTfh response in ageing was, however, associated with an enhanced pro-inflammatory gene expression signature, suggesting that excess inflammation can limit the response to vaccination. We were able to confirm that these enhanced inflammatory signatures associated with poor antibody titre in an independent cohort of influenza vaccinees. Together, this suggests that formation of antigen-specific Tfh cells is essential for high titre antibody responses, and that excessive T cell-extrinsic inflammatory factors contribute to poor cTfh cell and antibody responses to vaccination.

In order to understand what type of immune response supports protective antibody production upon vaccination we comprehensively analysed the immune response to seasonal influenza vaccination by measuring IgG responses to 32 HA proteins, 8 cytokines and chemokines, B cells, CD4^+^ T cells and HA-specific CD4^+^ T cells. This combined approach enabled us to identify which parameters were correlated with the antibody response after vaccination. In contrast to previous studies (*22, 37*), we observed that the frequencies of circulating B cell populations including plasmablasts did not correlate with the long-term antibody response. This suggests that the circulating plasmablasts observed seven days after vaccination in people are likely biomarkers of the early extrafollicular antibody response that provides a short initial burst of antibodies, rather than of long-lived plasma cells (*38*). However, the down-regulation of BAFF and up-regulation of one of its receptors, BCMA, in serum over the first 7 days after vaccination clearly demonstrate that dynamic regulation of B cell responses through BAFF and its receptors are tightly intertwined with the magnitude of the antibody output to vaccination. Consistent with this, the up-regulation of *TNFRSF17*, the gene for BCMA, has been well reported to correlate with vaccine titres in PBMC transcriptomics (*8, 39-41*). Importantly, we show that expansion of Tfh cells, both polyclonal and HA-specific, were consistently correlated with the magnitude of antibody responses across two cohorts. Through sequencing the HA-specific cells we were able to show that cTfh cells are recalled from the resting memory CD4^+^ T cell compartment, that public TCR clonotypes are readily detectable in antigen-specific cTfh cells, and that antigen-specific cTfh cells share a transcriptional program with lymph node Tfh that includes the downregulation of TNF and IL-2 signalling. As age is one of the key factors that influences antibody responses to vaccines (*42-44*), by clearly defining ideal immune responses in younger individuals we were able to gain novel insights into how ageing negatively impacts the immune response resulting in low titre antibody responses following seasonal influenza vaccination.

Impaired T cell responses to vaccines in ageing has been proposed to be caused by contraction of the TCR repertoire and the accumulation of terminally differentiated effector cells. However, in our study, we observed impaired antigen-specific Tfh differentiation in older people, despite no defect in the overall antigen-specific CD4^+^ T cell response. Through analysing the clonal relatedness in the TCR repertoire of HA peptide-specific T cells from before and after vaccination, we observed that similar frequencies of d7 cTfh cell clonotypes were recalled by vaccination from resting memory CD4^+^ T cell compartment in both age groups. This suggests that the ability of memory cells to be recalled by vaccines in ageing is not compromised. Furthermore, we observed no difference in the diversity of the TCR repertoire of the cTfh cells that formed after vaccination between age-groups. Therefore, this suggests that a loss of diversity in the T cell repertoire with age does not explain the impaired Tfh differentiation we observed in our study. Impaired Tfh responses to vaccines have been observed in humans and mice (*9, 45*), however, interestingly the formation of pre-Tfh cells remains intact but there is a failure to generate bona fide GC Tfh cells (*45, 46*). Together with our data, this suggests that instead of intrinsic defects in CD4^+^ T cells pre-vaccination in older people, the reduced cTfh cell frequency post vaccination may be explained by extrinsic factors, such as inflammation, that lead to a failure to appropriately acquire the GC Tfh cell gene signature.

Ageing is often associated with a state of low grade inflammation, termed ‘inflammaging’(*47-49*). However, in this study we observed no age-related difference in inflammatory gene signatures in HA-specific CD4^+^ T cells at day 0, which suggests the cTfh cell response was negatively impacted by inflammation post vaccination, rather than pre-existing inflammation having affected resting memory T cell function prior to vaccination. Because older people are more at risk of severe health outcomes after infection, significant effort has been made to alter vaccine formulations to enable them to be effective in this age group. Modifications to vaccines to increase the antibody response in older individuals now include increasing the antigen dose and using more potent adjuvants (*50, 51*), which enhance the inflammatory response. We have previously demonstrated in older people and aged mice that type 1 interferon and TLR7 signalling is important for conventional dendritic cells type 2 to support Tfh differentiation after vaccination (*9*). These studies, together with the data presented here, prompt the hypothesis that while some types inflammation are ‘good’ for promoting long-lived antibody responses in ageing, cytokines such as TNF and IL-2 are ‘bad’ and negatively impact a Tfh-supported response to vaccination. Consistent with this, inhibition of inflammatory monocyte recruitment into the skin of older people enhanced the local CD4 T cell response to varicella zoster virus antigen challenge (*52*). Therefore, vaccination strategies that support humoral immunity through limiting ‘bad’ pro-inflammatory signalling may be the key to improving vaccine efficacy in older people.

Our data demonstrates that IL-2 is one signalling pathway that is normally downregulated during Tfh differentiation. IL-2 is a cytokine produced by T cells early after T cell activation that promotes clonal expansion, and favours a Th1 cell fate at the expense of Tfh cell differentiation (*28, 31*), in line with the negative relationship observed here between cTfh cell differentiation and the response to IL-2 in those cells. Tfh cells are reported to be IL-2 producers as a consequence of heightened T cell receptor signalling, and whilst they typically do not respond to IL-2, they provide a source of this cytokine to support non-Tfh cells (*29*). Transcript levels for *IL2* were not increased in cTfh cells from older donors, however *IL2RB* was increased suggesting that the failure to downregulate an IL-2 gene signature in ageing could be due to increased response to IL-2 in the local environment after vaccination. Alternatively, IL-2 signalling gene signature may reflect a failure to fully acquire the Tfh transcriptional program, which may result from memory T cells receiving inadequate Tfh lineage commitment signals from the local environment, such as dendritic cell priming which is known to be impaired in aged mice (*9*).

TNF signalling in cTfh cells was also negatively correlated with Tfh differentiation and antibody response upon vaccination. While this cytokine is necessary for the formation of primary B cell follicles and follicular dendritic cell networks that underpin the formation of germinal centers (*53-56*), it has also been implicated in the loss of germinal centres and disorganisation of secondary lymphoid organ architecture during infections and immunizations in mice (*57, 58*). The role for TNF in Tfh cell differentiation or survival remains unclear, and it is noteworthy that this pleiotropic cytokine can regulate many aspects of T cell biology, including NF-κB signalling, TCR signalling, and apoptosis pathways (*59*). In our study, this TNF signalling signature did not include the *TNF* gene, nor were *TNF* transcripts altered in cTfh cells with age. While in our study, serum concentrations for both TNF and IL-2 were below the limit of detection in most samples, serum TNF levels and TNF production from memory B cells have been reported to increase with age (*60, 61*), suggesting our TNF signature in cTfh cells may result from paracrine sources within the lymph node. TNF/TNF-receptor signalling has been linked to reduced memory CD4^+^ T cell frequencies after influenza infection in mice (*62, 63*), and T cells from older people have been found to be more sensitive to TNF-alpha-induced apoptosis via the extrinsic pathway (*64, 65*), suggesting TNF may impact Tfh cell expansion or survival. In agreement with this, we observed some gene overlap between the apoptosis and TNF signatures downregulated in young cTfh cells on day 7 relative to day 0, and elevated in cTfh cells in older people. More research is required to understand how TNF may regulate germinal centre biology, particularly as elevated TNF correlated with impaired germinal centres and absence of Tfh cells observed in patients that succumbed to COVID-19 infection (*66, 67*), and has been negatively correlated with Tfh cell frequency in symptomatic SARS-COV2 infected patients (*68*). Therefore, while it is unclear whether TNF acts directly on T cells to impair Tfh differentiation or survival, or has a broader effect on lymphoid organ architecture, our data indicates that persistence of TNF signalling in cTfh cells is linked to poor vaccine responses.

The need to develop effective vaccines that are effective in all age groups has been emphasised by the COVID19 pandemic, in which older people are more likely to become seriously ill, or die, after infection(*69*). Promisingly, multiple SARS-CoV2 vaccines are able to induce strong antibody responses in older people. While a single dose of adenoviral vectored vaccines results in a lower antibody titre in older people, this can be boosted to comparable levels to younger adults with a second dose (*70, 71*). Mechanistic studies in mice show that this is due to the second vaccine dose enhancing Tfh cells and the germinal center response to a greater extent in aged mice than in younger adult animals (*72*). COVID19 mRNA vaccines induce high titre antibodies after two doses in all age groups (*73-75*), and generate superior germinal center responses than protein subunit vaccines in young mice (*76*) and humans (pre-printed in (*77*)). Together with the data presented here, this demonstrates that effective vaccines are ones that promote the germinal centre response. Therefore vaccination strategies that induce the optimal inflammatory environment to support Tfh differentiation is key to generating enduring antibody mediated immunity.

## Materials and Methods

### Human Cohorts

Peripheral blood was collected from healthy UK adults recruited through the NIHR Bioresource and vaccinated with the trivalent influenza vaccine. Participants were selected on the basis of having at least one allele of either *HLADR*0701* or *HLADR*1101* as determined by single nucleotide polymorphism typing using the UKBiobank v2.1 Axiom array. Two cohorts were collected: Cohort 1, northern hemisphere winter 2014-15, n=16 participants 18-36 years old, n=18 participants 66-75 years old; Cohort 2, northern hemisphere winter 2016-17, n=21 participants 18-36 years old, n=21 participants 66-98 years old. Venous blood was collected for serum and into EDTA coated tubes for peripheral blood mononuclear cell (PBMC) processing on the day of vaccination (prior to administration of the vaccine), 7 and 42 days after vaccination. PBMC were isolated by density gradient separation using Histopaque-1077 (Sigma), frozen in Foetal Bovine Serum supplemented with 10% Dimethyl sulfoxide (Sigma) and kept in liquid nitrogen prior to analysis by flow cytometry or flow sorting for RNA sequencing. Lymph node samples were taken from patients recruited from the renal transplant live donor program at Cambridge University Hospitals NHS Foundation Trust, as part of the routine operative procedure, as described previously (*20, 78*).

### Ethics

All human blood and tissue were collected in accordance with the latest revision of the Declaration of Helsinki and the Guidelines for Good Clinical Practice (ICH-GCP). The seasonal UK influenza vaccination cohort was collected with UK local research ethics committee approval (REC reference 14/SC/1077), using the facilities of the Cambridge Bioresource (REC reference 04/Q0108/44). Lymph node samples were collected from UK adults undergoing surgery for their own medical care, under ethical approval from UK Health Research Authority (REC reference 11/EE/0355, respectively), at Cambridge University Hospitals, and processed at the Babraham Institute. Written informed consent was received from all volunteers.

### Flu HA and cytokine Luminex

IgG to influenza HA proteins was measured before and after vaccination by Luminex using magnetic beads coated with full length recombinant haemagglutinin proteins, as previously reported (*79*). Proteins were expressed using baculovirus expression system in insect cells and coupled to Bio-plex Pro Magnetic COOH Beads (Bio-Rad, Hercules, CA). Titres are represented as arbitrary units per mL, calculated from a total IgG standard curve. The lower limit of detection for each HA protein was defined as the mean+ 2 standard deviations measured for the HA-coated beads with secondary antibody only. Where indicated, pre-existing IgG titres were subtracted to from day 7 and 42 titres to calculate vaccination-induced IgG responses. Serum samples were analysed for cytokine and chemokine concentrations using Human Magnetic Luminex kit (Cat# LXSAHM, R&D systems) custom ordered to detect the following analytes: BAFF, CXCL13, BCMA, APRIL, TWEAK, Osteopontin, SCF, Light. Analytes were included for further analysis if more than 50 percent of samples had values above the lower limit of detection.

### Hemagglutination inhibition assay

Antibody titers pre and post vaccination were determined using the hemagglutination inhibition (HAI) assay using the standard WHO protocol, as previously described (*80*). Sera were treated overnight with receptor-destroying enzyme (Denka Seiken Co.) and were subsequently tested by standard methods using 4 HA units of virus and a 0.5% suspension of turkey red blood cells. HAI titers were recorded as the reciprocal of the highest dilution of the serum that completely inhibited agglutination of erythrocytes by 4 HA units of the virus.

### Flow cytometry of B cells and FACS sorting of T lymphocytes

Cryopreserved mononuclear cells were thawed and rested for 1 hour at 37°C. Fc receptors were blocked using anti-human CD32 antibody (clone 6C4, eBioscience). 4 million PBMCs were stained with a panel of antibodies to measure B cells (Table S2), acquired on BD LSR Fortessa5 cytometer, and gated according to the strategy outlined in Fig. S1. For T cell staining and sorting, between 15-40 million PBMCs were first treated with 50nM dasatinib (Sigma) for 10minutes at 37°C, and then stained with PE-conjugated tetramers for 2 hours at room temperature with methods and reagents that have been previously reported (*15*) (Benaroya Research Institute Tetramer core facility). Tetramers were loaded with peptides of hemagglutinin (HA) protein specific to the A/California/04/2009(H1N1) influenza strain (GenBank: ACQ76318.1), with the HLADR*0701 tetramer containing the peptide ITFEATGNLVVPRYAFAMER which corresponds to amino acids 257-276, and HLADR*1101 tetramer contained the peptide FYKNLIWLVKKGNSYPKLSK which corresponds to amino acids 161-180(*15*). Tetramer stained PBMCs were then enriched for memory CD4^+^ T cells using by magnetic separation (MagniSort enrichment, eBioscience), and subsequently stained with the antibody panel outlined in Table S3. HA-specific T cells were isolated using a FACSAria™ Fusion Sorter (BD Biosciences) by first excluding unwanted cell types using a dump channel consisting of viability dye and antibodies to CD14, CD16, and CD19. CD3+CD4+CD45RA-tetramer binding cells were sorted and further phenotyping was performed using the gating strategy outlined in Fig. S2. Up to 1000 HA-specific tetramer+ cells were sorted directly into 9μL of lysis buffer containing RNAse inhibitor as per the SMART-Seq® v4 Ultra® Low Input RNA Kit for Sequencing manual (Takara), with the additional markers CXCR5 and PD1 used to isolate tetramer binding cTfh cells in d7 samples. Additional CD4+ T cell subsets were analysed post acquisition as per the gating strategy outlined in Fig. S3.

### RNA sequencing and data processing

mRNA was converted to cDNA using the SMART-Seq v4 Ultra Low Input RNA Kit (Takara Bio). Sequencing libraries were subsequently generated using the Nextera XT DNA Library Prep Kit (Illumina), followed by sequencing on the Illumina HiSeq 2000 with ∼50 million 100-bp single-end reads per sample. mRNA from lymph node CD4^+^CD45RA^−^ T cell populations was isolated from 1000 cells sorted into lysis buffer from 6 individuals as previously described (*20*). Sequencing reads were aligned to the reference human genome GRCh38 using HISAT2 (*81*), and quantitated using Rsubread package (*82*). Samples were excluded on the basis of poor cDNA quality prior to sequencing, or where drop-out genes with zero counts represented more than 80% of total reads. Genes were filtered based on having more than 10 reads in 20% of samples, yielding 19113 genes. Unwanted variation due to batch effects and sex was determined using the RUVseq package (*83*) for each cohort seperately using the model.matrix [∼data$sex +data$group], where the ‘group’ factor represents the combination of time-point, cell type and age-group. The RUVseq output was incorporated into DESeq2 model matrix design using the following model [design = ∼ *W*_1 + *W*_2 + *W*_3 + sex + group] where *W* represents the three RUVseq variance factors identified for each cohort. DESeq2 fold changes were adjusted using the lfcShrink normal method, and variance stabilised normalisation was applied to the counts to give an expression value per gene (DESeq2 package (*84, 85*)). Significantly differentially expressed genes had adjusted p-values of <0.1 and log2fold change of 0.5 or greater, with consistent changes across both cohorts or comparisons. Pathway analysis was performed using the Hallmark genesets (MSigDB collections (*27*)) on gene lists ranked by log2Fold change using the fgsea R package (*86*). Significantly enriched pathways had adjusted p-values of <0.1 and normalised enrichment scores of <-1 or >1, consistently across both cohorts or comparisons. The lymph node germinal centre Tfh signature was generated by comparing Tfh with CD4+CD45RA-CXCR5- and CD4+CD45RA-CXCR5+PD1-non-Tfh cell populations. Genes were selected on the basis of a DESeq2 adjusted P-value cutoff <0.1 and log2 fold change of greater of less than 0.5, for both non-Tfh populations. Pathways analysis using the fgsea package was used to define enriched pathways as described above.

Signature genes were determined from the fgsea defined leading-edge gene lists for TNF signalling pathway, IL-2 STAT5 signalling pathway, and inflammation pathways (Interferon gamma, Inflammatory response, and Complement combined). Genes present in the leading-edge for both cohorts were included, and where a gene was present across multiple pathways, it was assigned to the pathway where it was ranked highest, ensuring each gene signature was non-overlapping. VSN normalised gene expression values for selected signatures were then converted to z-scores with all time-points combined, and then z-scores were summed for each sample (*87*). Microarray data of PBMCs from GSE74813 (*8, 88*) were re-analysed as follows: Raw. CEL files were downloaded from GEO with the corresponding annotation data. CEL files were read into R via Affy package(*89*) and were normalised using VSN (*84*). The dataset was then filtered to select only individuals with paired day 0 and day 7 samples. Gene z-scores were then calculated and summed across gene signatures for each sample as described above, with the summed gene signature scores for d0 subtracted from d7 scores for each sample for correlation to antibody titres, which were made available upon request (*8*).

### T cell receptor sequencing and clonotype analysis

TCRβ clonotypes were called from adaptor-trimmed RNA sequencing fastq files using MIXCR (version 2.1.9;(*90*)) run in RNA-Seqmode with rescuing of partial alignments and set to collate *TCRA* or *TCRB* clonotypes at the amino acid level and requiring more than five reads to identify a clonotype. Clonotype diversity was determined using vdjtools (version 1.0.3; (*91*)). Recalled *TCRB* clones were determined by analysing paired d0 and d7 samples from the same individual for both cohorts combined, and clonotype defined by CDR3 amino acid sequence, V, D and J gene usage. Public *TCRB* clonotypes were analysed for both cohorts combined, and each time-point separately, with V and J gene usage among public clones compared for each time-point and genotype separately.

### Vaccination trajectory and pseudotime analysis

The trajectory analysis was assembled as previously described (*7, 92*). Briefly, the frequencies for immune parameters were first normalized for abundance by subtracting the mean and dividing by the standard deviation for both cohorts combined. Variables with missing values were excluded, leaving 35 variables analysed for each time-point (Table S1) Principal components analysis was then performed, in which the samples from different time-points showed separated by PC1. Therefore, the immune parameters that had a correlation to PC1 greater than 0.4 were included (*n* = 23 cell types). The diffusion maps algorithm was then applied to the scaled frequencies using the destiny R package(*93, 94*), the resulting diffusion pseudotime values (described as vaccination trajectory) were scaled to a range of 0 to 1 and compared between time-points and age-groups. Correlations between vaccination trajectory and cell type frequencies were analysed by Spearman’s correlation.

### Statistics

All statistical tests for cell type frequencies assumed nonparametric data. Fold change for immune parameters was calculated by dividing d7 or d42 frequency by d0, and post vaccination increases relative to baseline were determined by subtracting d0 from d7 or d42 data. Two-group comparisons were made using either two-tailed Mann-Whitney tests or Wilcoxon tests for paired data from the same individual at different time points. Multiple group comparisons p-values were calculated using Dunn’s post hoc test. Correlation analyses used Spearman correlation, with the exception of log normalised TCRB clone frequency analysis which used Pearson correlation. As necessary, FDR (Benjamini-Hochberg) adjustment was used to adjust for multiple testing on two-group comparisons. Heatmaps of manual gated cell subsets that were altered by vaccination or age group were generated using Pheatmap package (*95*).

## Data Availability

The authors confirm that the data supporting the findings of this study are available within the article [and/or] its supplementary materials.

## Acknowledgements

The NIHR Cambridge Biomedical Research Center (BRC) is a partnership between Cambridge University Hospitals NHS Foundation Trust and the University of Cambridge, funded by the National Institute for Health Research (NIHR). We are indebted to the NIHR Cambridge BRC volunteers for their participation and we thank the NIHR Cambridge BRC staff for their contribution in co-ordinating the vaccinations and venepuncture. We thank the staff of the Babraham Institute Flow Cytometry Facility and the Sequencing Facility for their technical assistance. We are grateful to the Babraham Bioinformatics Group for their help. This study was supported by H2020 European Research Council funding awarded to MAL (637801-TWILIGHT) and the Biotechnology and Biological Sciences Research Council (BBS/E/B/000C0427, BBS/E/B/000C0428, and the Campus Capability Core Grant to the Babraham Institute). DLH is supported by a National Health and Medical Research Council Australia Early-Career Fellowship (APP1139911). JCL is supported by a Wellcome Trust Intermediate Clinical Fellowship (105920/Z/14/Z). MAL is an EMBO Young Investigator and Lister Institute Prize Fellow. This paper presents independent research supported by the NIHR Cambridge BRC. The views expressed are those of the authors and not necessarily those of the NIHR or the Department of Health and Social Care.

## Author contributions

Conceptualization, M.A.L., E.J.C and D.L.H; Methodology, D.L.H, E.J.C., S.I., J.W., M.Z., E.A.J., W.W.K and M.A.L; Investigation, D.L.H, E.J.C., S.I., J.W., M.Z., and M.A.L; Clinical Leads, J.C.L and E.J.C. Writing – Original Draft Preparation, D.L.H., and M.A.L.; Writing – Review and Editing, all authors; Project Administration, M.A.L., D.L.H and E.J.C; Funding Acquisition, M.A.L.

## Supp Figures

Figure S1: B cell flow cytometry gating strategy

Figure S2: CD4+ T cell flow cytometry gating strategy

Figure S3: HA-specific CD4+ T cell sorting strategy

Figure S4: Cytokine and CD4+ T cell variables altered after vaccination

Figure S5: CXCR3 and CCR6 expression on HA-specific CD4+ T cells

Figure S6: Cell number and clonotype number from T cell sequencing

Figure S7: Age-related differences in cytokines and HA-specific CD4^+^ T cell parameters

Table S1: Immunological variables

Table S2: Inflammation, TNF and IL-2 gene signatures

Table S3: Antibody panel for B cells

Table S4: Antibody panel for T cells

Data File S1: 18-36 year old samples d7 cTfh vs d0 gene list

Data File S2: Lymph Node Germinal center Tfh gene signature

Data File S3: d7 Tfh gene set

Data File S4: d7 Vaccination genes

Data File S5: Ageing d7 cTfh vs d0 gene list

