## Supplementary materials for "Impaired HA-specific T follicular helper cell and antibody responses to influenza vaccination are linked to inflammation in humans"

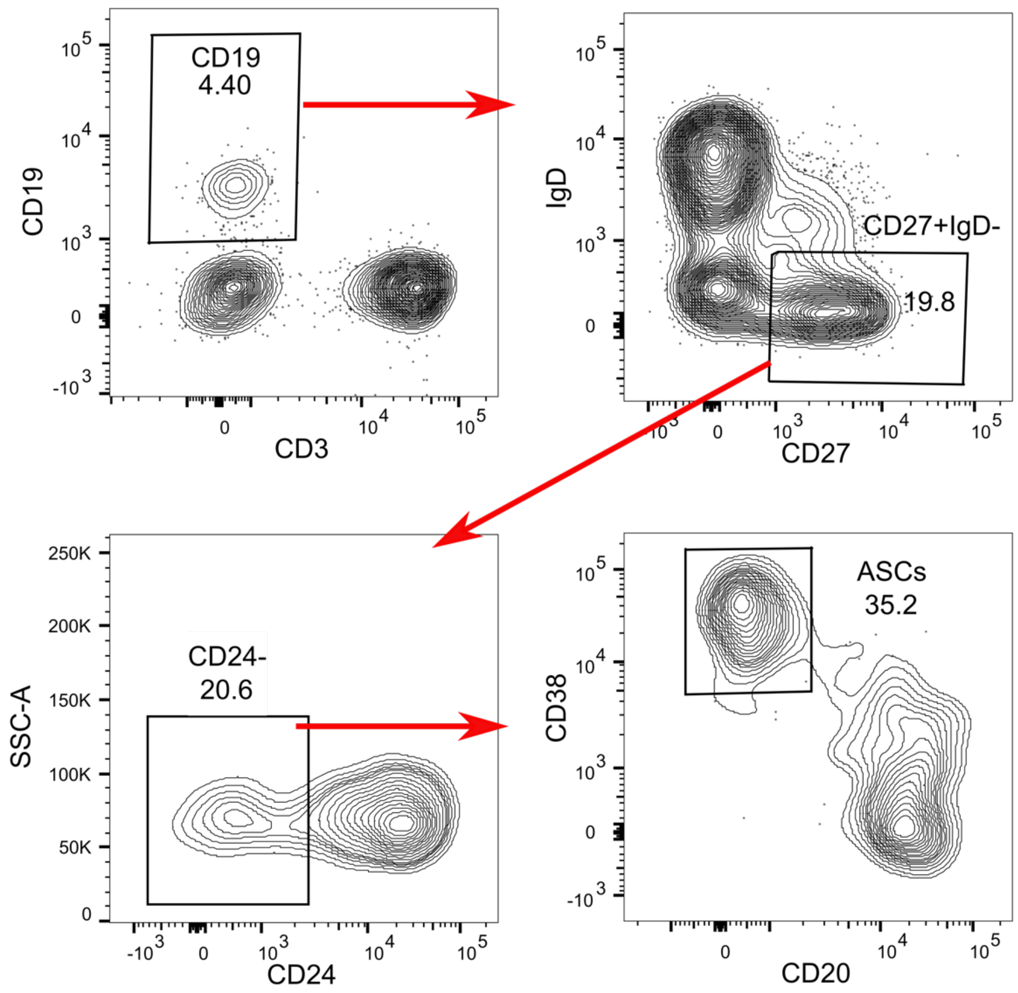


**Fig. S1: B cell flow cytometry gating strategy**

Representative flow cytometry plots for B cell immunophenotyping pre-gated on single, live lymphocytes, stained with the panel outlined in Table S3.

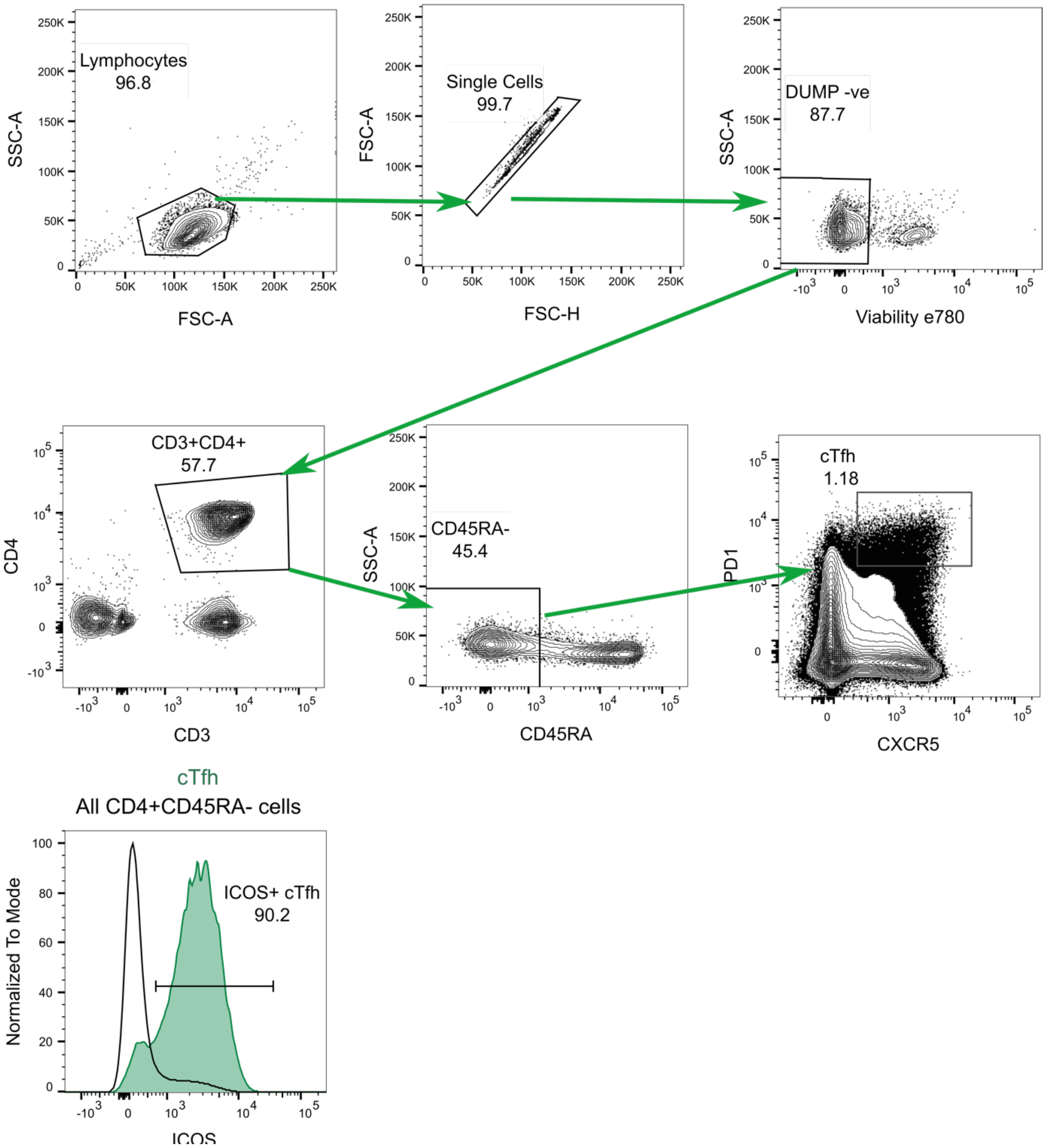


**Fig. S2: CD4+ T cell flow cytometry gating strategy**

Representative flow cytometry plots for the analysis of CD4+ T cells, stained with the panel outlined in Table S4. Histograms show representative staining for ICOS expression on d7 cTfh cells (green) shown relative to the entire CD4+CD45RA- T cell population (black outline) from the same sample.

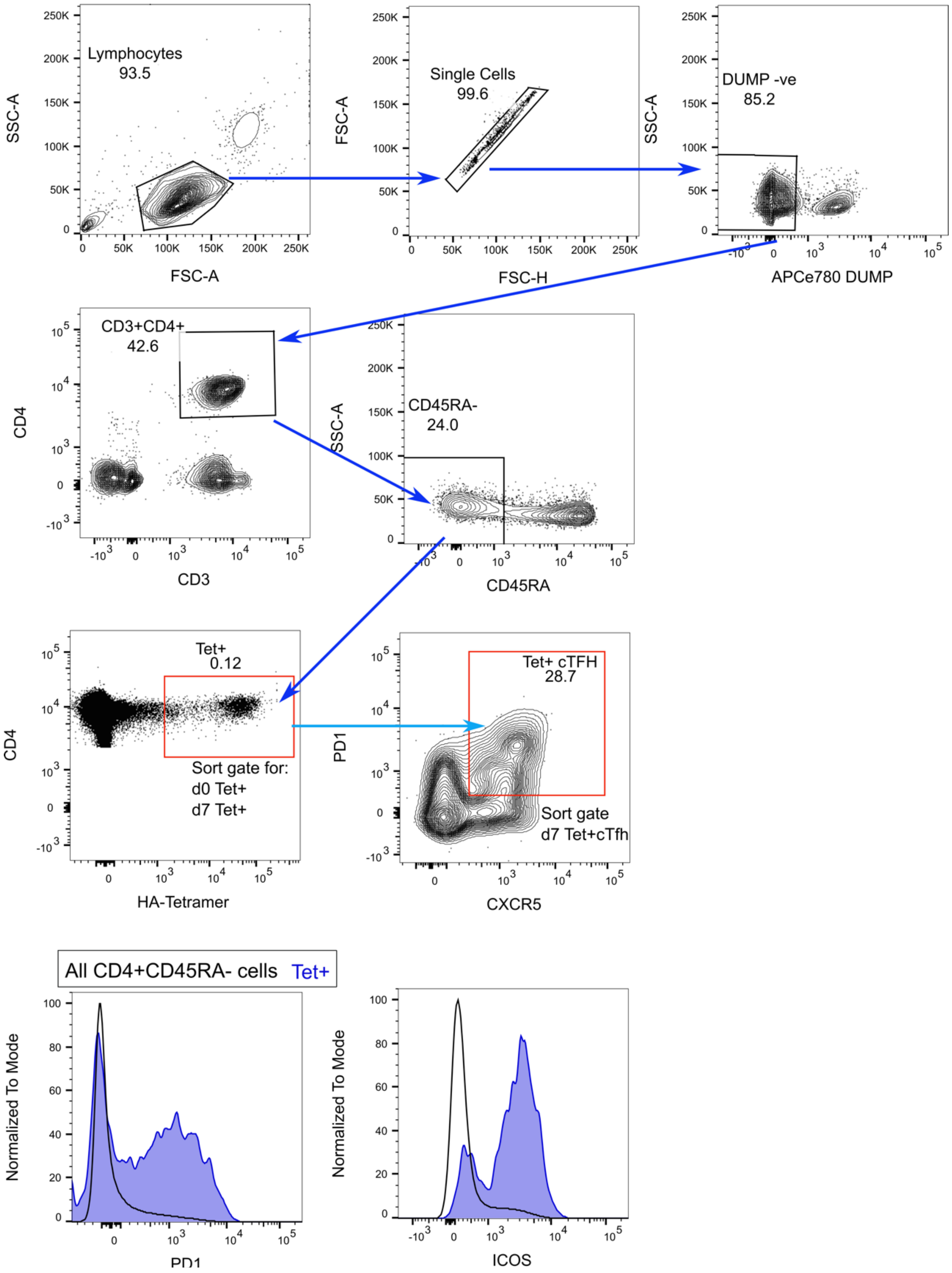


**Fig. S3: HA-specific CD4+ T cell sorting strategy**

Representative flow cytometry plots for the cell sorting of HA-specific Tet+ cells, stained with the panel outlined in Table S4. Red gates indicate the sorted populations on d0 and/or d7. Histograms show representative staining for PD1 and ICOS expression on d7 Tet+ cells (blue) shown relative to the entire CD4+CD45RA- T cell population (black outline) from the same sample.

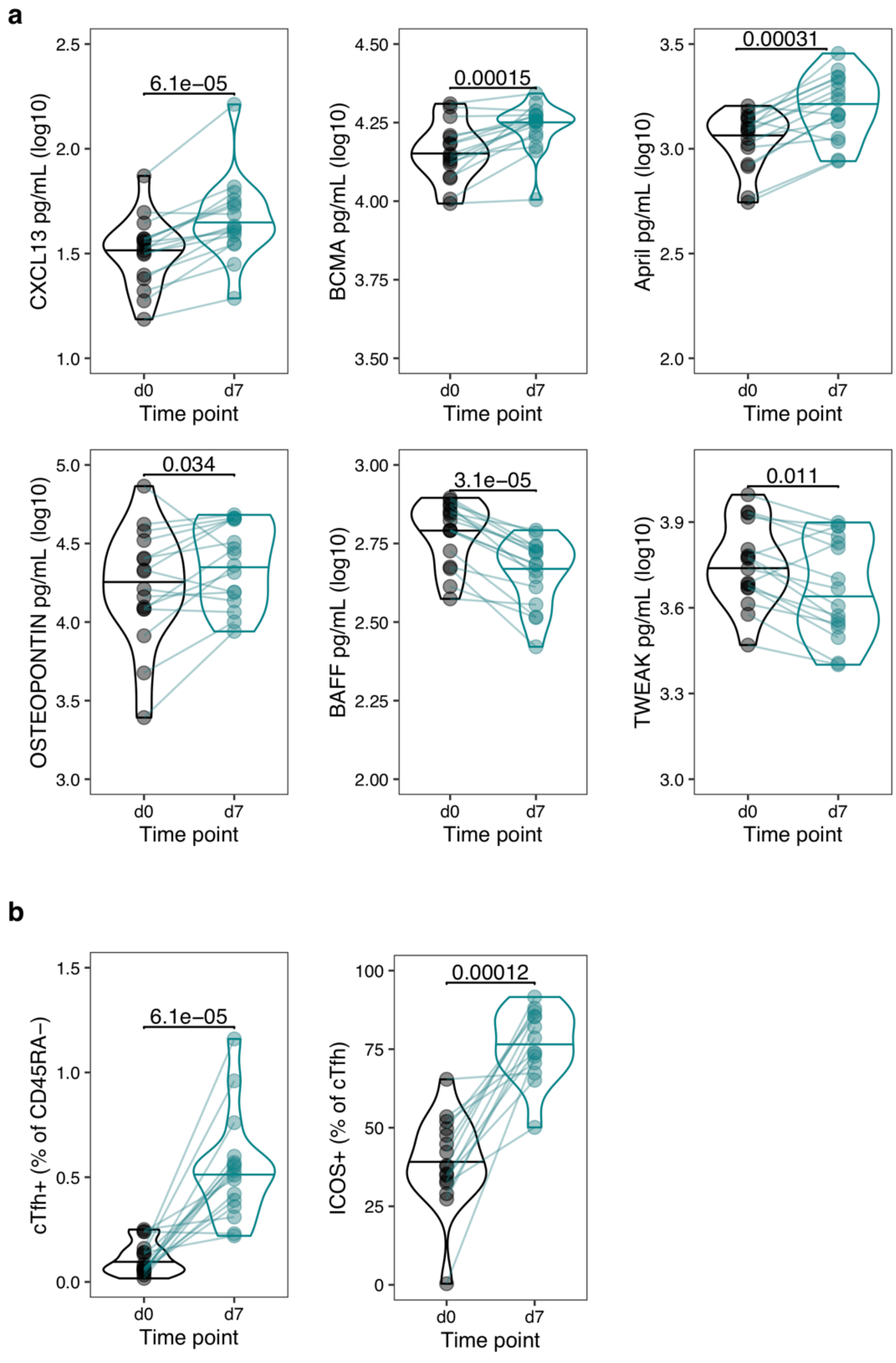


**Figure S4: Cytokine and CD4+ T cell variables altered after vaccination in 18-36 year old individuals**

A) The concentration of cytokines in serum (log10 pg/mL) at d0 and d7. Variables with significant $\log_2$ fold change in Fig. 1 shown. B) The percentage of cTfh+ cells among CD45RA- T cells, and ICOS+ cells among cTfh cells at d0 and d7. Cohort 1 only (n=16). Paired p-values determined using Wilcoxon signed rank-test.

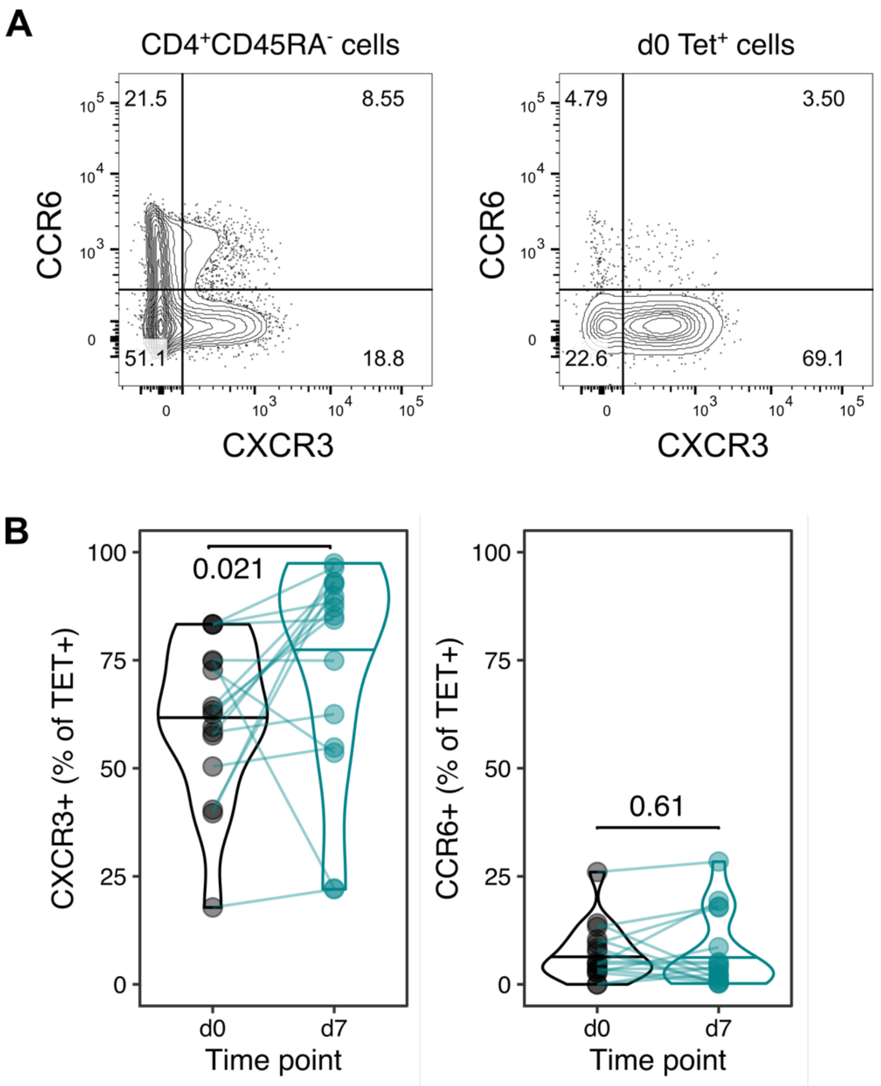


**Fig. S5: CXCR3 and CCR6 expression on HA-specific CD4+ T cells**

A) Representative flow cytometry plots for CXCR3 and CCR6 expression on CD4+CD45RA- T cells and HA-specific Tet+ cells. B) The quantification of CXCR3 and CCR6 expression on Tet+ cells on day 0 and d7. Paired p-values determined using Wilcoxon signed rank-test.

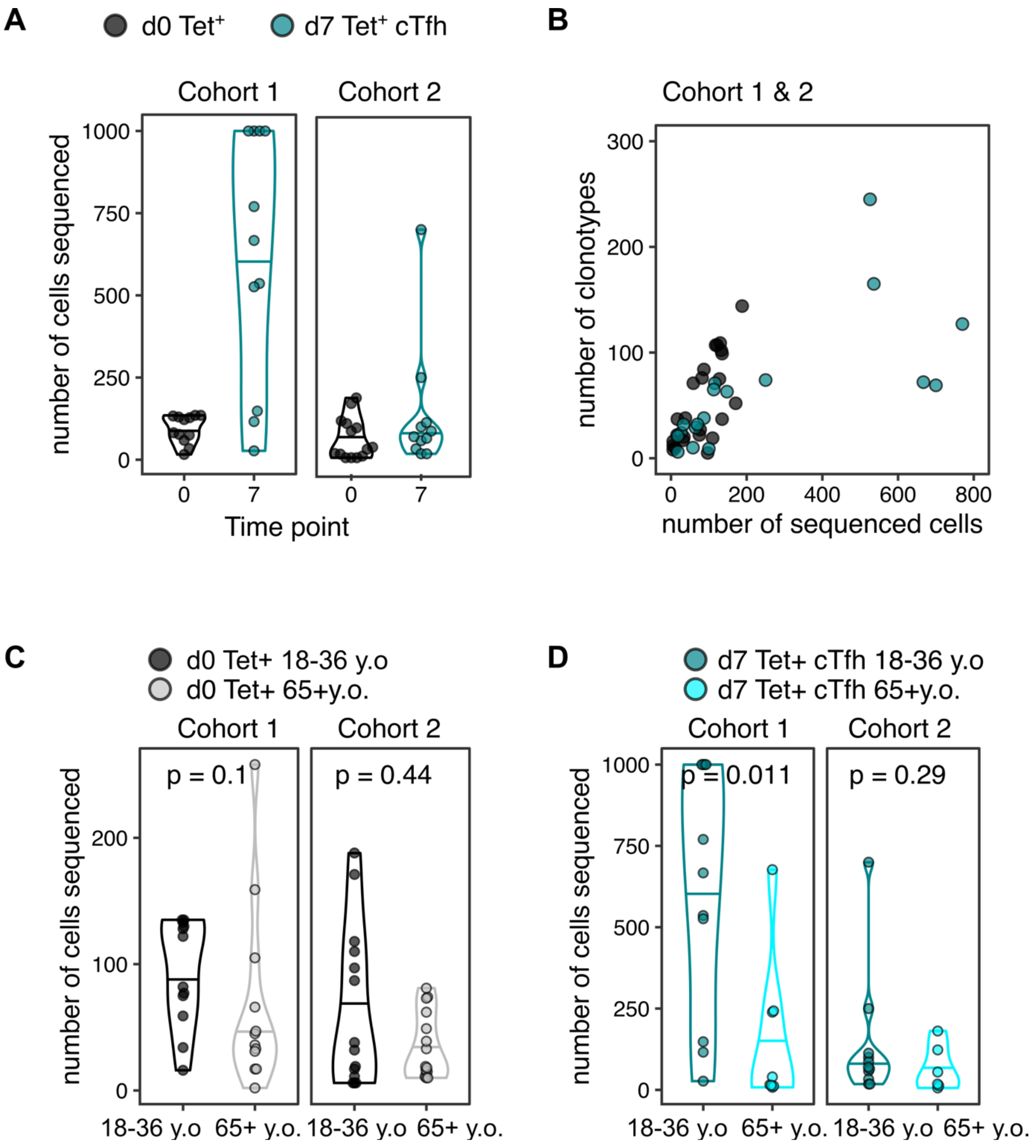


**Fig. S6: Cell number and clonotype number from T cell sequencing**

A) The number of cells sorted and sequenced at each time-point for each cohort, 18-36 y.o samples only. B) The number of sequenced cells displayed against the number of *TCRB* clonotypes recovered from sequencing, for 18-36 year old samples from both cohorts combined. C) The number of Tet+ cells sorted and sequenced at d0 for each cohort and for each age group. D) The number of Tet+cTfh cells sorted and sequenced at d7 for each cohort and for each age group. P-values calculated by Mann-Whitney *U* test.

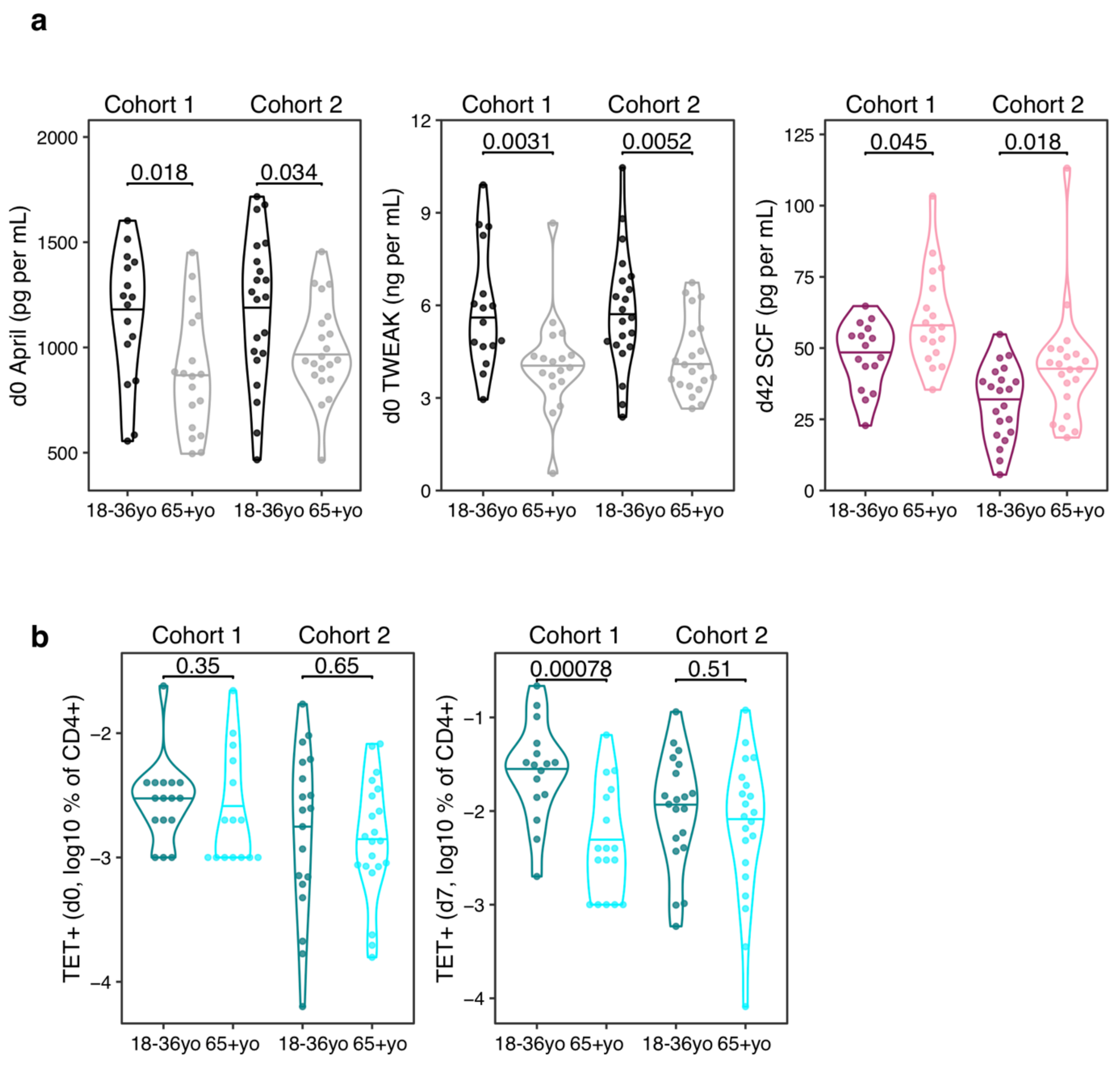


**Figure S7: Age-related differences in cytokines and HA-specific CD4$^{+}$ T cell parameters.**

A) The serum concentration (log10 pg/mL) of d0 April, d0 TWEAK, and d42 SCF across age-groups and cohorts. Variables with consistently significant (padj <0.05) across both cohorts as in Fig. 5G shown. B) The percentage of Tet+ cells among CD4+ T cells at d0 and d7 across age-groups and cohorts as in Fig. 5H. Within-cohort age group differences were determined using the Mann-Whitney *U* test.

### Table S1: Immunological variables

| Var # | var name | included in initial trajectory analysis | retained post PC1 trimming |
|---|---|---|---|
| 1 | cytokine_Luminex_d0_April..pg.mL. | Y | Y |
| 2 | cytokine_Luminex_d0_BAFF..pg.mL. | Y | |
| 3 | cytokine_Luminex_d0_BCMA..pg.mL. | Y | |
| 4 | cytokine_Luminex_d0_CXCL13..pg.mL. | | |
| 5 | cytokine_Luminex_d0_Light..pg.mL. | Y | |
| 6 | cytokine_Luminex_d0_OSTEOPONTIN..pg.mL. | Y | |
| 7 | cytokine_Luminex_d0_SCF..pg.mL. | Y | |
| 8 | cytokine_Luminex_d0_TWEAK..pg.mL. | Y | |
| 9 | cytokine_Luminex_d7_April..pg.mL. | Y | |
| 10 | cytokine_Luminex_d7_BAFF..pg.mL. | Y | |
| 11 | cytokine_Luminex_d7_BCMA..pg.mL. | Y | |
| 12 | cytokine_Luminex_d7_CXCL13..pg.mL. | | |
| 13 | cytokine_Luminex_d7_Light..pg.mL. | Y | |
| 14 | cytokine_Luminex_d7_OSTEOPONTIN..pg.mL. | Y | |
| 15 | cytokine_Luminex_d7_SCF..pg.mL. | Y | |
| 16 | cytokine_Luminex_d7_TWEAK..pg.mL. | Y | |
| 17 | cytokine_Luminex_d42_April..pg.mL. | | |
| 18 | cytokine_Luminex_d42_BAFF..pg.mL. | | |
| 19 | cytokine_Luminex_d42_BCMA..pg.mL. | | |
| 20 | cytokine_Luminex_d42_CXCL13..pg.mL. | | |
| 21 | cytokine_Luminex_d42_Light..pg.mL. | | |
| 22 | cytokine_Luminex_d42_OSTEOPONTIN..pg.mL. | | |
| 23 | cytokine_Luminex_d42_SCF..pg.mL. | | |
| 24 | cytokine_Luminex_d42_TWEAK..pg.mL. | | |
| 25 | flu_Ab_Luminex_d0_A.BJ03 | | |
| 26 | flu_Ab_Luminex_d0_A.Cali09 | Y | Y |
| 27 | flu_Ab_Luminex_d0_A.gfHK99 | | |
| 28 | flu_Ab_Luminex_d0_A.HK68 | Y | |
| 29 | flu_Ab_Luminex_d0_A.Jap57 | Y | |
| 30 | flu_Ab_Luminex_d0_A.NewCal99 | Y | Y |
| 31 | flu_Ab_Luminex_d0_A.PC73 | Y | Y |
| 32 | flu_Ab_Luminex_d0_A.Perth09 | Y | Y |
| 33 | flu_Ab_Luminex_d0_A.PR8 | | |
| 34 | flu_Ab_Luminex_d0_A.rheaNC93 | Y | Y |
| 35 | flu_Ab_Luminex_d0_A.SC18 | Y | Y |
| 36 | flu_Ab_Luminex_d0_A.SH13 | | |
| 37 | flu_Ab_Luminex_d0_A.Swi13 | Y | Y |
| 38 | flu_Ab_Luminex_d0_A.SZ06 | | |
| 39 | flu_Ab_Luminex_d0_A.Taiw13 | Y | Y |
| 40 | flu_Ab_Luminex_d0_A.Tex12 | Y | Y |
| 41 | flu_Ab_Luminex_d0_A.Tex91 | | |
| 42 | flu_Ab_Luminex_d0_A.USSR77 | | |
| 43 | flu_Ab_Luminex_d0_A.Vic11 | Y | Y |
| 44 | flu_Ab_Luminex_d0_A.Viet04 | | |
| 45 | flu_Ab_Luminex_d0_B.Bris08 | Y | Y |
| 46 | flu_Ab_Luminex_d0_B.Maly04 | Y | Y |
| 47 | flu_Ab_Luminex_d0_B.Mass12 | Y | Y |
| 48 | flu_Ab_Luminex_d0_B.Phu13 | Y | Y |
| 49 | flu_Ab_Luminex_d0_B.Wis10 | Y | Y |
| 50 | flu_Ab_Luminex_d0_cH4.7 | Y | |
| 51 | flu_Ab_Luminex_d0_cH5.1Cal | | |
| 52 | flu_Ab_Luminex_d0_cH5.1PR8 | | |
| 53 | flu_Ab_Luminex_d0_cH5.3 | Y | |
| 54 | flu_Ab_Luminex_d0_cH9.1 | | |
| 55 | flu_Ab_Luminex_d0_H5.Head | | |
| 56 | flu_Ab_Luminex_d0_H9.Head | | |
| 57 | flu_Ab_Luminex_d7_A.BJ03 | | |
| 58 | flu_Ab_Luminex_d7_A.Cali09 | Y | Y |
| 59 | flu_Ab_Luminex_d7_A.gfHK99 | | |
| 60 | flu_Ab_Luminex_d7_A.HK68 | Y | |
| 61 | flu_Ab_Luminex_d7_A.Jap57 | Y | |
| 62 | flu_Ab_Luminex_d7_A.NewCal99 | Y | Y |
| 63 | flu_Ab_Luminex_d7_A.PC73 | Y | Y |
| 64 | flu_Ab_Luminex_d7_A.Perth09 | Y | Y |
| 65 | flu_Ab_Luminex_d7_A.PR8 | | |
| 66 | flu_Ab_Luminex_d7_A.rheaNC93 | Y | Y |
| 67 | flu_Ab_Luminex_d7_A.SC18 | Y | Y |
| 68 | flu_Ab_Luminex_d7_A.SH13 | | |
| 69 | flu_Ab_Luminex_d7_A.Swi13 | Y | Y |
| 70 | flu_Ab_Luminex_d7_A.SZ06 | | |
| 71 | flu_Ab_Luminex_d7_A.Taiw13 | Y | Y |
| 72 | flu_Ab_Luminex_d7_A.Tex12 | Y | Y |

| | | | |
|---|---|---|---|
| 73 | flu_Ab_Luminex_d7_A.Tex91 | | |
| 74 | flu_Ab_Luminex_d7_A.USSR77 | | |
| 75 | flu_Ab_Luminex_d7_A.Vic11 | Y | Y |
| 76 | flu_Ab_Luminex_d7_A.Viet04 | | |
| 77 | flu_Ab_Luminex_d7_B.Bris08 | Y | Y |
| 78 | flu_Ab_Luminex_d7_B.Maly04 | Y | Y |
| 79 | flu_Ab_Luminex_d7_B.Mass12 | Y | Y |
| 80 | flu_Ab_Luminex_d7_B.Phu13 | Y | Y |
| 81 | flu_Ab_Luminex_d7_B.Wis10 | Y | Y |
| 82 | flu_Ab_Luminex_d7_cH4.7 | Y | |
| 83 | flu_Ab_Luminex_d7_cH5.1Cal | | |
| 84 | flu_Ab_Luminex_d7_cH5.1PR8 | | |
| 85 | flu_Ab_Luminex_d7_cH5.3 | Y | |
| 86 | flu_Ab_Luminex_d7_cH9.1 | | |
| 87 | flu_Ab_Luminex_d7_H5.Head | | |
| 88 | flu_Ab_Luminex_d7_H9.Head | | |
| 89 | flu_Ab_Luminex_d42_A.BJ03 | | |
| 90 | flu_Ab_Luminex_d42_A.Cali09 | | |
| 91 | flu_Ab_Luminex_d42_A.gfHK99 | | |
| 92 | flu_Ab_Luminex_d42_A.HK68 | | |
| 93 | flu_Ab_Luminex_d42_A.Jap57 | | |
| 94 | flu_Ab_Luminex_d42_A.NewCal99 | | |
| 95 | flu_Ab_Luminex_d42_A.PC73 | | |
| 96 | flu_Ab_Luminex_d42_A.Perth09 | | |
| 97 | flu_Ab_Luminex_d42_A.PR8 | | |
| 98 | flu_Ab_Luminex_d42_A.rheaNC93 | | |
| 99 | flu_Ab_Luminex_d42_A.SC18 | | |
| 100 | flu_Ab_Luminex_d42_A.SH13 | | |
| 101 | flu_Ab_Luminex_d42_A.Swi13 | | |
| 102 | flu_Ab_Luminex_d42_A.SZ06 | | |
| 103 | flu_Ab_Luminex_d42_A.Taiw13 | | |
| 104 | flu_Ab_Luminex_d42_A.Tex12 | | |
| 105 | flu_Ab_Luminex_d42_A.Tex91 | | |
| 106 | flu_Ab_Luminex_d42_A.USSR77 | | |
| 107 | flu_Ab_Luminex_d42_A.Vic11 | | |
| 108 | flu_Ab_Luminex_d42_A.Viet04 | | |
| 109 | flu_Ab_Luminex_d42_B.Bris08 | | |
| 110 | flu_Ab_Luminex_d42_B.Maly04 | | |
| 111 | flu_Ab_Luminex_d42_B.Mass12 | | |
| 112 | flu_Ab_Luminex_d42_B.Phu13 | | |
| 113 | flu_Ab_Luminex_d42_B.Wis10 | | |
| 114 | flu_Ab_Luminex_d42_cH4.7 | | |
| 115 | flu_Ab_Luminex_d42_cH5.1Cal | | |
| 116 | flu_Ab_Luminex_d42_cH5.1PR8 | | |
| 117 | flu_Ab_Luminex_d42_cH5.3 | | |
| 118 | flu_Ab_Luminex_d42_cH9.1 | | |
| 119 | flu_Ab_Luminex_d42_H5.Head | | |
| 120 | flu_Ab_Luminex_d42_H9.Head | | |
| 121 | HAI_d0_log2.titre | Y | Y |
| 122 | HAI_d7_log2.titre | Y | Y |
| 123 | HAI_d42_log2.titre | | |
| 124 | B_flow_d0_ASCs...Freq..of.CD19 | | |
| 125 | B_flow_d0_CD19.ve...Freq..of.Parent | | |
| 126 | B_flow_d0_CD27.ve...Freq..of.CD19 | | |
| 127 | B_flow_d7_ASCs...Freq..of.CD19 | | |
| 128 | B_flow_d7_CD19.ve...Freq..of.Parent | | |
| 129 | B_flow_d7_CD27.ve...Freq..of.CD19 | | |
| 130 | T_flow_d0_CTFH.HIGH.HIGH.OF.CD45RA | Y | Y |
| 131 | T_flow_d0_ICOS.OF.CTFH.HIGH.HIGH | Y | Y |
| 132 | T_flow_d0_CD45RA.OF.CD3.CD4 | Y | |
| 133 | T_flow_d0_CD3.CD4.OF.LIVE | Y | |
| 134 | T_flow_d0_CTFH.OF.TET.ALL | Y | Y |
| 135 | T_flow_d0_TET.ALL.OF.CD4 | Y | Y |
| 136 | T_flow_d0_ICOS.TET.ALL.OF.CD45RA | Y | Y |
| 137 | T_flow_d0_PD1.OF.TET.ALL | Y | Y |
| 138 | T_flow_d0_ICOS.OF.TET.ALL | | |
| 139 | T_flow_d7_CTFH.HIGH.HIGH.OF.CD45RA | Y | Y |
| 140 | T_flow_d7_ICOS.OF.CTFH.HIGH.HIGH | Y | Y |
| 141 | T_flow_d7_CD45RA.OF.CD3.CD4 | Y | |
| 142 | T_flow_d7_CD3.CD4.OF.LIVE | Y | |
| 143 | T_flow_d7_CTFH.OF.TET.ALL | Y | Y |
| 144 | T_flow_d7_TET.ALL.OF.CD4 | Y | Y |
| 145 | T_flow_d7_ICOS.TET.ALL.OF.CD45RA | Y | Y |
| 146 | T_flow_d7_PD1.OF.TET.ALL | Y | Y |
| 147 | T_flow_d7_ICOS.OF.TET.ALL | | |

**Table S2: Inflammation, TNF and IL-2 gene signatures**

| Inflammation score | TNF Score | IL2 Score |
|---|---|---|
| ADM | AHNAK | CDKN1A |
| CCL2 | ARL4A" | CEBPB |
| CCL20 | BHLHE40 | CEBPD |
| CCL5 | CAPG | DUSP1 |
| CD36 | EMP1 | DUSP2 |
| CD55 | FAH | FOS |
| CD74 | GADD45B | G0S2 |
| CD82 | HOPX | GCH1 |
| CFH | IFNGR1 | ID2 |
| CSF2RB | KLF6 | IER2 |
| CTSH | PNP | IER3 |
| CXCL8 | PTCH1 | IER5 |
| DPP4 | RGS16 | JUN |
| EREG | RNH1 | JUNB |
| FCER1G | RORA | KDM6B |
| FCN1 | SLC1A5 | KLF10 |
| FPR1 | SLC39A8 | MSC |
| GNAI3 | SYNGR2 | NFKB2 |
| GP1BA | XBP1 | NFKBIA |
| HLA-B | | NINJ1 |
| HLA-DRB1 | | PHLDA2 |
| ICAM1 | | PTGS2 |
| IL18R1 | | RELB |
| IL2RB | | SERPINB2 |
| IL6 | | TANK |
| ITGA5 | | TNFAIP3 |
| LAP3 | | ZC3H12A |
| LCK | | ZFP36 |
| LCP2 | | |
| LY6E | | |
| MYD88 | | |
| NFKB1 | | |
| NLRP3 | | |
| OLR1 | | |
| OSM | | |
| PDE4B | | |
| PLAUR | | |
| RCE1 | | |
| RHOG | | |
| RIPK1 | | |
| RNF4 | | |
| S100A12 | | |
| SERPING1 | | |
| SLAMF7 | | |
| SRI | | |
| STAT4 | | |
| STX4 | | |
| TIMP1 | | |
| TNFAIP6 | | |
| VCPIP1 | | |

**Table S3: Antibody panel for B cells**

| Marker | clone | Fluorochrome | company | catalog no. |
|---|---|---|---|---|
| viability dye | n/a | eFluor780 | Thermo Fisher | 65-0865-18 |
| CD14 | 61D3 | APC-eF780 | Thermo Fisher | 47-0149-42 |
| CD16 | eBioCB16 | APC-eF780 | Thermo Fisher | 47-0168-42 |
| CD3 | UCHT1 | BV605 | Biolegend | 300460 |
| CD19 | HIB19 | BB515 | BD Biosciences | 564456 |
| IgD | IA6-2 | BV421 | BD Biosciences | 563813 |
| CD38 | HIT2 | APC | Thermo Fisher | 17-0389-42 |
| CD20 | 2H7 | PECY7 | Biolegend | 302312 |
| CD27 | M-T271 | BV650 | BD Biosciences | 564894 |
| CD24 | eBioSN3 | PerCP-eFluor 710 | Thermo Fisher | 46-0247-42 |

**Table S4: Antibody panel for T cells**

| Marker | clone | Fluorochrome | company | catalog no. |
|---|---|---|---|---|
| viability dye | n/a | eFluor780 | Thermo Fisher | 65-0865-18 |
| CD14 | 61D3 | APC-eF780 | Thermo Fisher | 47-0149-42 |
| CD16 | eBioCB16 | APC-eF780 | Thermo Fisher | 47-0168-42 |
| CD19 | HIB19 | APC-eF780 | Thermo Fisher | 47-0199-42 |
| CD3 | UCHT1 | BUV 395 | BD Biosciences | 563546 |
| CD45RA | HI100 | BUV737 | BD Biosciences | 564442 |
| CD4 | RPA-T4 | PercpCy5.5 | BD Biosciences | 560650 |
| CXCR5 | RF8B2 | BB515 | BD Biosciences | 564624 |
| PD1 | eBioJ105 | APC | Thermo Fisher | 17-2799-42 |
| ICOS | ISA-3 | biotin | Thermo Fisher | 13-9948-82 |
| CXCR3 | 1C6/CXCR3 | BV421 | BD Biosciences | 562558 |
| CCR6 | 11A9 | BV786 | BD Biosciences | 563704 |
| Streptavidin | - | BV650 | BD Biosciences | 563855 |
